# Multi-ancestry genetic architecture of sleep duration and its relationship to other sleep and psychiatric phenotypes

**DOI:** 10.1101/2025.05.19.25327902

**Authors:** Isabelle Austin-Zimmerman, Daniel F. Levey, Joseph D. Deak, Marco Galimberti, Keyrun Adhikari, Jonathan R. I. Coleman, the VA Million Veteran Program, Daniel J. Buysse, Peter W.F. Wilson, Tamar Sofer, J. Michael Gaziano, Daniel J. Gottlieb, Murray B. Stein, Marta Di Forti, Joel Gelernter

## Abstract

Differences in sleep duration, quality, and timing are associated with variation in cognition, health outcomes, and quality of life. Genetic studies may help explain the underlying mechanisms of sleep and its relationships to other conditions.

Our previous work highlighted risk loci associated with short (<6hrs) and long sleep (>9hrs), using data from the UK Biobank and the Million Veteran Program. We build on this work by conducting a genome wide association study (GWAS) and multi-ancestry meta-analysis of sleep duration as a quantitative trait. We used LD score regression (LDSC) to evaluate the correlation between sleep duration and other traits, and genomic structural equation modelling (genomicSEM) to consider the relationships between traits of interest.

We identify 234 independent genome-wide significant loci for sleep duration, of which 143 are novel. The average impact of each risk variant amounts to approximately ±0.86minutes (sd=0.19), with a sum total of ± 220.5 minutes across all genome-wide significant loci. We support previous findings showing the most strongly associated gene is *PAX8*. Linkage disequilibrium score regression shows that the genetic architecture of sleep duration is largely distinct from other measures of sleep quality and sleep disorders. We see several examples of negative correlation between deleterious traits and the quantitative measure of sleep duration reported here, contrasting with positive associations with long and short sleep (e.g., depression, ADHD, cannabis use disorder, smoking). We derive genomic-SEM models that show short and long sleep load on separate factors, as does overall sleep duration loading alone.

This is the largest available GWAS of sleep duration, and the first to extend analyses outside of European ancestry populations. We identify novel loci for sleep duration and provide insight to the shared and unique genetic architecture across multiple sleep and neuropsychiatric traits.

## Introduction

Sleep is essential to health, and poor sleep is associated with a range of negative health outcomes, as well as all-cause mortality^1^. ‘Good’ sleep can be measured in various ways – sleep quality, timing, and efficiency all have impacts on overall health, as does sleep duration^1^. The extent to which poor sleep directly causes these negative health consequences is unclear, though recent work employing Mendelian randomisation analysis is providing insight to the causality of these observed associations^2–4^.

The genetic architecture of sleep duration has been studied in several recent genome-wide association studies (GWAS). Work by our group and others has identified several well-replicated genetic associations, including variants mapping to the *VRK2*, *TCF4*, *FOXP2*, and *PAX8* genes^5–9^. These studies demonstrate shared genetic architecture between sleep duration and a range of cardiometabolic, psychiatric, and cognitive traits. The SNP-based heritability for overall sleep duration is reported to be 9.8% from the most recent GWAS^6^, but estimates from twin studies range from 30-45%^9–12^, indicating that there is more to discover at the level of genetic variation.

Our previous study of long and short sleep demonstrated a modest positive genetic correlation between the two traits (Rg=0.16)^5^. Despite this observation, there was also a similarity in the associations with other phenotypes – where both long and short sleep were positively correlated to a range of comorbid conditions including major depressive disorder (MDD), cannabis use disorder (CUD), and post-traumatic stress disorder (PTSD), among others^5^. These findings support clinical, epidemiological, and genetic studies in indicating that the relationship between sleep duration and negative health outcomes is non-linear; both too little and too much sleep can be linked to morbidity.

Here, we build on previous work^5^ to investigate the genetic architecture of sleep duration further. We present the largest GWAS on sleep duration conducted thus far, including 648,125 participants of diverse ancestry from the UK Biobank (UKB) and Million Veteran Program (MVP). We consider sleep duration as a quantitative measure and investigate the relationship between overall sleep duration and other sleep and neuropsychiatric phenotypes.

## Methods

### Participants

Details on the recruitment and overall sample for the UKB are described in ^13^ for UKB, and in ^14^ for MVP. The UKB study was approved by the North-West Research Ethics Committee (ref 06/MREC08/65) in accordance with the Declaration of Helsinki. Research involving the MVP in general is approved by the VA Central Institutional Review Board. All participants in both cohorts provided written informed consent, and our included sample was reviewed to remove data for those participants who withdrew their consent.

### Genotype and phenotype measures

Full details on the genotyping, imputation and quality control measures applied to the genetic data have been described^13,14^. The fully imputed genetic data for UKB were made available in March 2018, and accessed under application 82087. This study includes MVP data release 4, made available in January 2021. For both cohorts, additional local post-imputation quality control procedures were applied. Participants were stratified into broad population groups (EUR and AFR included here) using principal component analysis^5^, using eigensoft^15^ for MVP and PC-AiR for UKB^16,17^. All SNPs with an imputation INFO score <0.6 or a minor allele frequency (MAF) <0.01, were removed. This was assessed within each ancestry group to avoid unnecessary filtration of variants with differing frequency by population. Participants with discordant self-report and genetically inferred sex were excluded due to risk of sample processing errors. Those with excessive genetic relatedness (more than 10 third-degree relatives within the respective cohorts) were removed. In addition, one of each pair of closely related individuals (closer than 2^nd^ degree relatives) were removed at random.

Both UKB and MVP recorded data on usual sleep duration at baseline. The phrasing of the question differed slightly, but in both cases the emphasis was on capturing hours of sleeping in a typical 24-hour period (UKB: “About how many hours sleep do you get in every 24h? (Please include naps)”; MVP: “How many hours do you usually sleep each day (24-hour period)?”. For UKB, the answers could be given as 1-23 hours, and participants who reported sleeping less than three hours or more than 12 were prompted to confirm their answers. Responses were given in hour increments. For MVP, the response options were multiple choice: 5 or less, 6, 7, 8, 9 or 10 or more.

### Genome-wide association study and meta-analyses

We downloaded the summary statistics from a recent GWAS of sleep duration in European-ancestry (EUR) participants from the UKB^13^. We used PLINK 2.0 to conduct GWAS using linear regression for EUR and African ancestry (AFR) subjects from the Million Veteran Program. The outcome variable was sleep duration (hours) and all analyses were adjusted for age, sex, 10 principal components, and in the case of UKB, genotype array^5,18^.

In the EUR sample, we ran a further analysis of sleep restricted to those who report sleeping between six and nine hours, to capture variation within the normal range, and consider how this compares to the full range of sleep duration, as well as to our prior binary analyses on short (<6 hours) and long (>9 hours) compared to healthy (7-8 hours) sleep^5^. This was run in UKB and MVP using the same protocol described for the main MVP sleep duration GWAS above.

The genetic correlation between the UK Biobank and MVP for self-reported sleep duration is 0.93. We used METAL^19^ to conduct inverse standard error weighted fixed effect meta-analyses within and across each ancestry groups. Within-ancestry summary statistics were filtered to remove any SNP that was not present in both UKB and MVP. The cross-ancestry summary statistics were filtered to include only variants that appeared at least once in both population groups across UKB and MVP data (but variants appearing in only one of the two cohorts were included if both ancestries were represented). Independent GWAS signals were identified through clumping of variants within a 3 megabase window and with an r^2^ threshold of 0.1.

### Post-GWAS analyses

#### Functional annotation and pathway-based tests

We used the Functional Mapping and Annotation GWAS platform (FUMA) version 1.5.2^20^ to annotate SNPs using positional mapping. We used reference panels from the relevant 1000 genome project data for AFR, AMR, EAS, and EUR populations. We used the associated platform Multi-marker Analysis of GenoMic Annotation (MAGMA) version 1.08^21^, using Ensembl build 85 to match SNPs to genes. We apply a Bonferroni corrected significance threshold of 0.05 divided by the included number of protein-coding genes (N=18,775-19,123). We also used MAGMA and PASCAL (Pathway SCoring Algorithm)^22^ to conduct genetic pathway and gene set analysis. These use different methods to assign pathway scores, all based on GWAS summary statistics. For PASCAL results, we applied a Bonferroni-corrected significance threshold for the 1,077 pathways tested of 4.5×10^-5^. MAGMA uses several pathway databases, which means many pathways are overlapping or very similar. Given the non-independent nature of these tests we applied a Benjamini-Hochberg false discovery rate (FDR) test.

We conducted SNP-level fine mapping using PolyFun (POLYgenic FUNctionally-informed fine-mapping)^23^ and SuSiE (Sum of Single Effects), as described previously^5^. We filtered variants with a posterior inclusion probability (PIP) score of 0.50 for nominally causative and 0.95 for likely causative.

We used the LDLink R package to create a list of all LD-proxy SNPs for our lead hits (defined as r^2^>0.8). We cross-checked these with previously published risk loci for sleep duration from references ^6^ and ^9^ to determine novelty. We used publicly available summary statistics to determine which of our lead SNPs or their LD-proxies had previously been linked to other sleep related traits (insomnia, chronotype, daytime napping, self-reported tiredness, accelerometer derived sleep duration).

#### Transcriptome wide association study

We used FUSION^24^ to conduct a transcriptome-wide association study (TWAS). This approach combines gene-expression data with the sleep duration GWAS summary statistics to identify patterns of gene expression. We used GTEx v8 multi-tissue expression weights from 49 tissues for expression imputation across all autosomes. We used the ‘FUSION.post_process.R’ script provided by the FUSION package (described in http://gusevlab.org/projects/fusion/)^24^ to define conditionally independent genes. We applied a multiple testing correction (as described in ^5^) based on 49 tissues and 27,977 Ensembl Gene IDs (compassing genes, non-coding transcripts, and pseudogenes) resulting in a p-value threshold of ≤ 3.65×10^-8^ (0.05/(49*27,977). If we identified a gene significantly expressed in two tissues, we report the association with the lower p-value.

#### Single cell polygenic regression

We employed the pathway-based polygenic regression method, scPagwas^25^, a single-cell enrichment method, to identify trait-relevant cell subpopulations by combining scRNA-seq data with GWAS summary statistics. This tool calculates a trait-relevant score (TRS) and p-values for each cell, which are then used to derive TRS and p-values for cell types. Significant TRS were defined at p-value<0.005 (corrected for 10 biological systems). For scRNA-seq data, we used datasets of 10 biological systems, which included nine organoids (brain, lung, intestine, heart, eye, liver, pancreas, kidney, and skin) and the immune system.

#### Global and local genetic correlation analyses

We calculated SNP-based heritability for sleep duration within each population group. We used linkage disequilibrium score regression (LDSC) with 1000 Genomes LD scores^26^ for EUR data, and sample-derived LD scores for the AFR data (described in^5^ and ^27^, https://github.com/immunogenomics/cov-ldsc).

We calculated the cross-population genetic correlation for sleep duration using Popcorn (version 0.9.6: https://github.com/brielin/Popcorn) for the cross-population analyses^28^. We assessed further genetic correlations between sleep duration and a range of sleep traits, including our data on short sleep (<6 hours), long sleep (>9 hours), overall sleep duration and normal range sleep duration (6-9 hours), plus neuropsychiatric, cardiometabolic, and sociodemographic traits (for full list of traits see supplementary data) using the EUR only data and LDSC^29–31^. We used the jackknife script within LDSC to assess the differences in correlation patterns between overall sleep, short sleep, long sleep, and normal range sleep duration.

We used SUPERGNOVA to identify the specific genomic regions driving the observed genetic correlation between six sleep traits (overall sleep duration, short sleep, long sleep, normal range sleep duration, insomnia^8^, and chronotype). Supergnova splits the genome into 2,353 ‘approximately independent regions’^33^ and estimates the genetic correlation at this localised level. We defined regions of significant local correlation according to a Bonferroni adjusted threshold p<2.1e-4 (0.05/2,353 regions).

#### Genomic Structural Equation Modelling

We conducted genomic structural equation modelling (gSEM) to identify common factors between 15 traits of interest^34^. These were the six sleep traits described above, along with alcohol consumption^35^, bipolar disorder^36^, cannabis use disorder (CUD)^37^, major depressive disorder (MDD)^38^, problematic alcohol use (PAU)^39^, physical activity^40^, PTSD^41^, schizophrenia^42^, subjective well-being^43^, and Townsend deprivation index^44^. We performed exploratory factor analysis (EFA) and confirmatory factor analysis (CFA) of these 15 genetically correlated traits. We considered multiple factor structures from one to six factors. EFA models were required to have a minimum of sum of squared (SS) loading ≥1 and variance explained by each factor >10%. Models meeting these criteria were compared based on cumulative variance explained by the overall factor structure. Where factor loadings were ≥0.20, traits were assessed for optimal CFA model fit according to chi-squared, standardised root mean square residual (SRMR), Akaike Information Criteria (AIC), and comparative fit index (CFI)^34^. We estimated CFA models using diagonally weighted least squares estimation and a smoothed genetic covariance matrix. This analysis was conducted in EUR data only and used the 1000 genome reference panel.

## Results

### Sample

We include a total of 646,218 individuals in our genome-wide meta-analysis of quantitative sleep duration (Table 1). The average sleep duration was similar in both studies, 7.2 hours (SD 1.1) in UKB and 7.0 hours (SD 1.3) in MVP. Table 1 summarises the age, ancestry, and sex distribution in the two cohorts. Although both are adult cohorts of majority EUR ancestry, the age, sex, and ancestry distributions differ between the two cohorts. The UKB recruited only adults aged 40-70, and the median age is 58. MVP recruits all adults over the age of 18, but median age is higher (66 years). UKB has a higher proportion of women compared to men. MVP has a greater proportion of non-EUR participants.

**Table 1.** Sample demographics.

|  | UKB | Million Veteran Program |
| --- | --- | --- |
| **Sample size *N*** | 453,506 | 192,712 |
| **Age mean years (*SD*)** | 56.8 (8.0) | 66.8 (11.6) |
| *median years* | 58 | 67 |
| **Sex (%female)** | 54.2 | 7.1 |
| **Genetic ancestry *N* (%total)** |  |  |
| EUR | 446,118 (98.4) | 163,536 (84.9) |
| AFR | 7,388 (1.6) | 29,176 (15.1) |
| **Sleep duration mean hours (*SD*)** | 7.2 (1.1) | 7.0 (1.3) |
| *median hours* | 7 | 7 |

### Genome-wide association study meta-analyses

In our EUR only meta-analysis we identify 153 independent genome-wide significant associations for sleep duration across 100 genomic risk loci. The lead hit (p=2.26×10^-66^) is rs62158206, which is in the *PAX8* region and has been previously associated with sleep duration in both overlapping and independent cohorts. This was also the strongest association with long sleep in our previous study. We identify 208 genes through the gene-based test, reaching a Bonferroni-corrected significance threshold of 2.68×10^-6^. The strongest association is *SGCZ* on chromosome 8, which is a protein-coding gene previously associated with several sleep traits as well as other psychiatric and musculoskeletal traits. We did not identify any genome-wide significant associations in our AFR-only meta-analysis of quantitative sleep duration. For more detail on the ancestry-specific analyses, see supplementary figures 3-8 and supplementary tables 1,2.

The cross-population meta-analysis identified 234 lead SNPs across 103 genomic risk loci (figure 1, supplementary table 3). Of these, 138 have never previously been associated with sleep duration (neither the specific locus nor any LD proxy SNP) (supplementary figure 2, supplementary tables 4,5), although nine of these have been associated with other sleep traits including insomnia (rs144938821, rs6711037, rs28420942, rs6561715, rs11039216^45^, rs6855246^8^), chronotype (rs4958316^32^), and short sleep (rs6855246, rs7313797, rs11689042^5^). The top association was within the same gene as in the EUR analysis, *PAX8\**rs2863957. Input SNPs were mapped to 19,019 protein coding genes, resulting in 291 significantly associated genes (p<2.63×10^-6^). The top gene here was *FOXP2*, previously associated with sleep duration in addition to many other traits.

**Figure 1.**
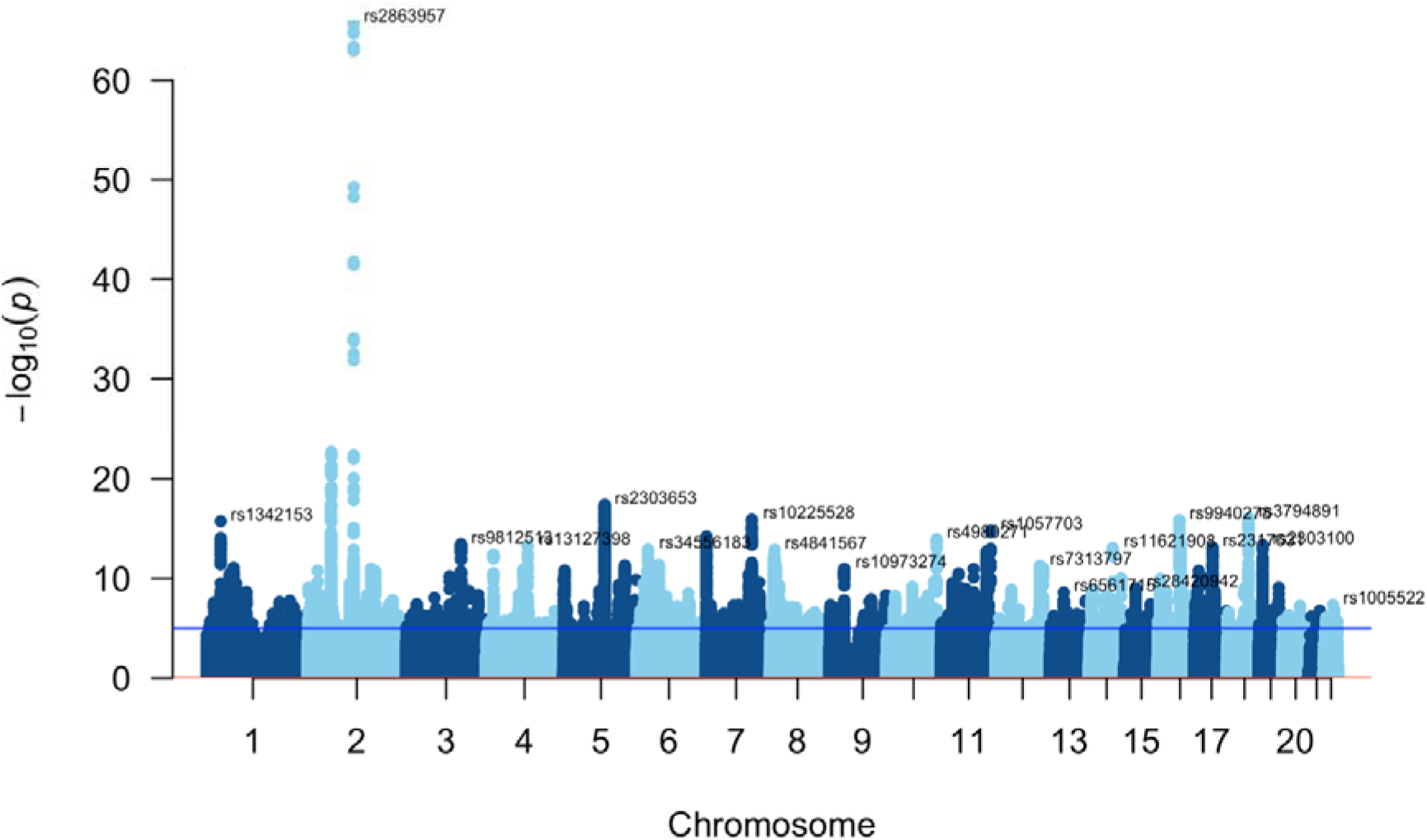
Manhattan plot showing the results for the cross-population meta-analysis of quantitative sleep duration in the UKB and MVP samples.

The effect size for these 234 lead SNPs equate to an impact on sleep duration ranging from a decrease of-2.29 minutes to an increase of 2.62 minutes, with the mean impact of each associated SNP being ±0.94 minutes. The cumulative effect of lead SNPs that increased sleep duration was 99 minutes, and the cumulative effect of lead SNPs that decreased sleep duration was-119 minutes. The variant having the strongest association with increasing sleep duration was *IGKV1OR2-108\**rs144938821 on chromosome 2 (beta= 0.044, SD= 0.004, p= 8.9×10^-23^; 2.6±0.24 minutes), which has a positive effect across MVP EUR, UKB EUR and UKB AFR but was not present in MVP AFR after filtering for minor allele frequency. The variant having the strongest association with decreasing sleep duration was *KMT2A\**rs7951019 on chromosome 11 (beta=-0.038, SD= 0.0059, p= 8.8×10^-1^^1^; - 2.3±0.35 minutes). This variant had a negative effect across both EUR cohorts, a positive effect in UKB AFR and was not present in MVP AFR.

### Pathway analysis

We conducted genetic pathway analysis using MAGMA and PASCAL. A total of 10 pathways reached significance in the MAGMA analysis (see supplementary table 6, supplementary figure 9). The PASCAL analysis revealed three pathways significantly associated with overall sleep duration following a Bonferroni correction for 1,077 pathways tested. Two of these were from the Reactome database (‘neuronal system’ and ‘transmission across chemical synapses’), and ‘long term depression’ from the KEGG database (see supplementary table 7).

### Transcriptome wide associations study

We conducted a TWAS and identified 171 conditionally independent genes (supplementary table 7, supplementary figures 10,11). Of these, 38 associations were identified across 13 brain tissues, with the remaining 133 associations spread across 34 tissues (Figure 3). A total of 70 of these conditionally independent genes were also identified in the gene-based GWAS test (Figure 2). The strongest TWAS genes that also appear in the gene-based test are *FTO* (TWAS Z-score=-8.7), *PPIP5K2* (TWAS Z=-8.6), *GIN1* (TWAS Z=8.0), *FOXP2* (TWAS Z=7.9), and *PAM* (TWAS Z=7.4).

**Figure 2.**
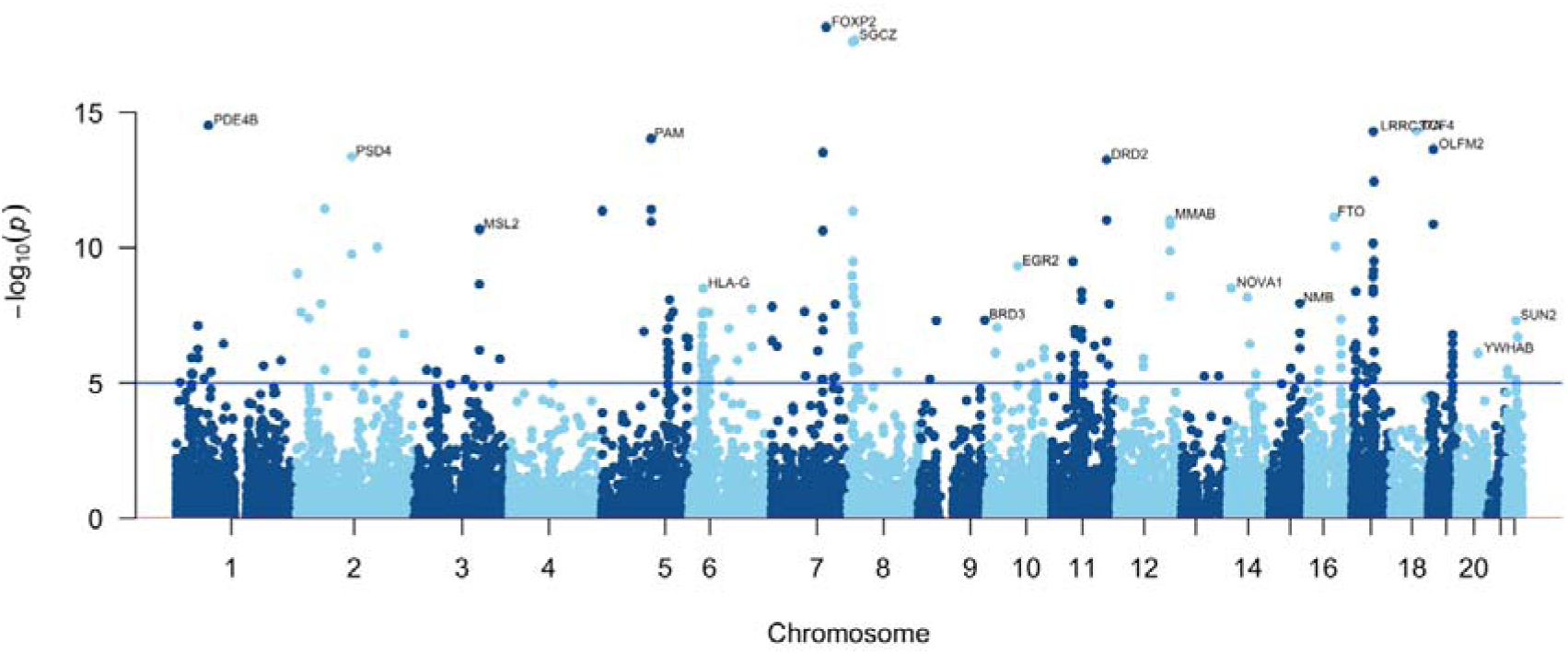
Gene-based Manhattan plot showing the results for the cross-population meta-analysis of quantitative sleep duration in the UKB and MVP samples.

**Figure 3.**
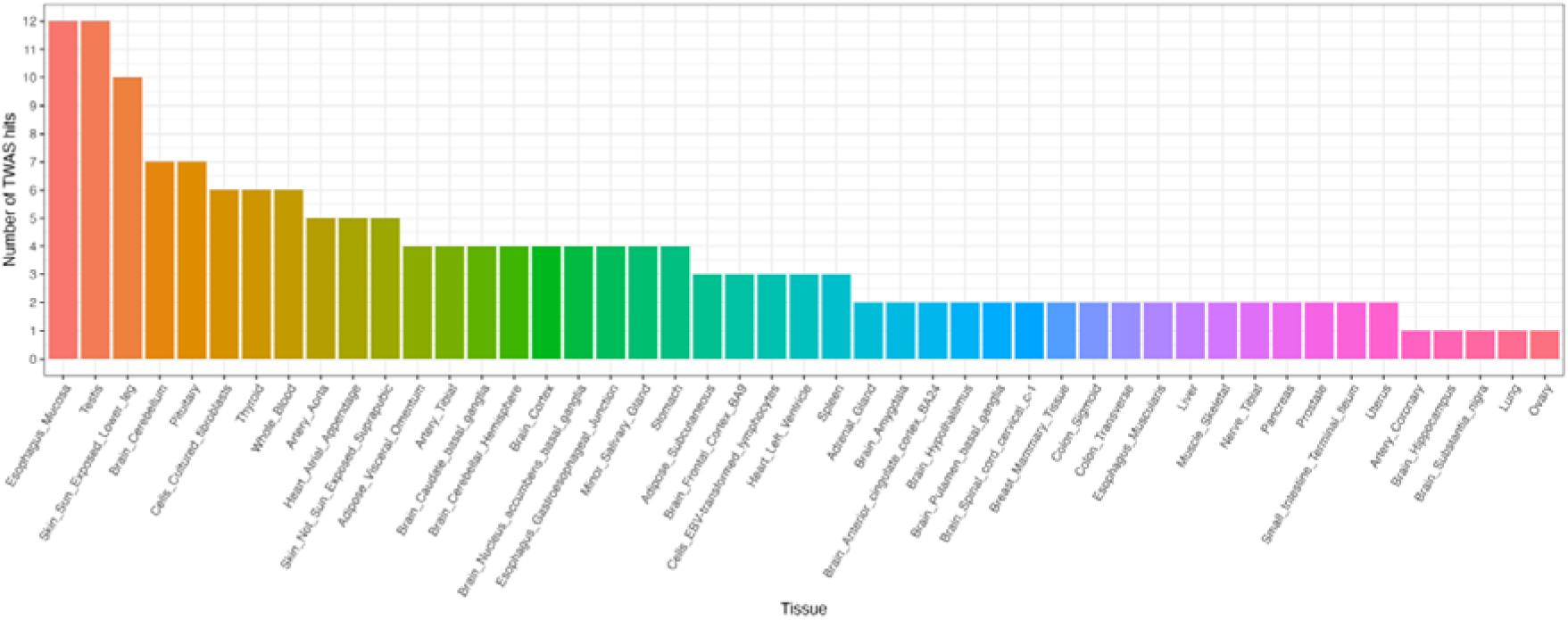
Frequency of TWAS hits across 47 tissues

### Single cell polygenic regression analysis

Our single-cell analyses in EUR data showed significant associations between sleep duration and cell types of each analyzed biological system (supplementary figures 12-21, supplementary table 8). The highest distribution of TRS was observed for the endothelial cells present in lung (Figure S-lung), which also showed the strongest association (*p*-value=2.49×10^-13^). This was followed by another lung cell type, mesenchymal (*p*-value=3.96 x10^-12^). The third strongest association was seen for cholangiocyte cells (*p*-value=1.68 x10^-11^) located in liver (Figure S-liver). The next strongest significant associations were with several cells belonging to the immune system (CD8.N, CD4.N2, CD4.N1, CD4.M, CD8.CM), the brain (excitatory neuron), and the eye (amacrine cell).

### Shared genetic architecture with other sleep traits

We conducted LDSC to test for the genetic correlation between overall sleep duration, sleep duration within the 6-9 hour range, our previously described short and long sleep traits, and other published sleep duration traits: sleep duration in children, self-reported under sleeper, self-reported over sleeper, self-reported tiredness, insomnia, and chronotype (morning person or not) (Figure 4, supplementary table 9). We identify the highest correlations with the under sleeper and over sleeper traits, in the expected direction (i.e. high positive correlation between short sleep and under sleeper (rg=0.93, sd=0.04, p=1.1 x10^-104^), high negative correlation between overall sleep duration and under sleeper (rg=-0.89, sd=0.04, p=1.0 x10^-140^); high positive correlation between over sleeper and both overall sleep duration (rg= 0.64, sd=0.07, p=6.9 x10^-19^) and long sleep (rg=0.883, sd=0.11, p=5.7 x10^-15^). We observe a low negative genetic correlation between chronotype (morningness) and sleep duration (rg=-0.12, sd=0.03, p=1.0 x10^-4^), suggesting that, on average, variants associated with overall sleep duration have the opposite effects in people with morning or evening chronotype. We see a low positive correlation between short sleep and chronotype (rg=0.11, sd=0.03, p=5.0 x10^-4^). As we previously reported, both long and short sleep are positively associated with insomnia (long sleep rg=0.25, sd=0.05, p=1.3 x10^-7^; short sleep rg=0.7, sd=0.03, p=4.8 x10^-169^). Overall sleep duration was negatively correlated with insomnia (rg=-0.48, sd=0.02, p=1.0 x10^-85^).

**Figure 4.**
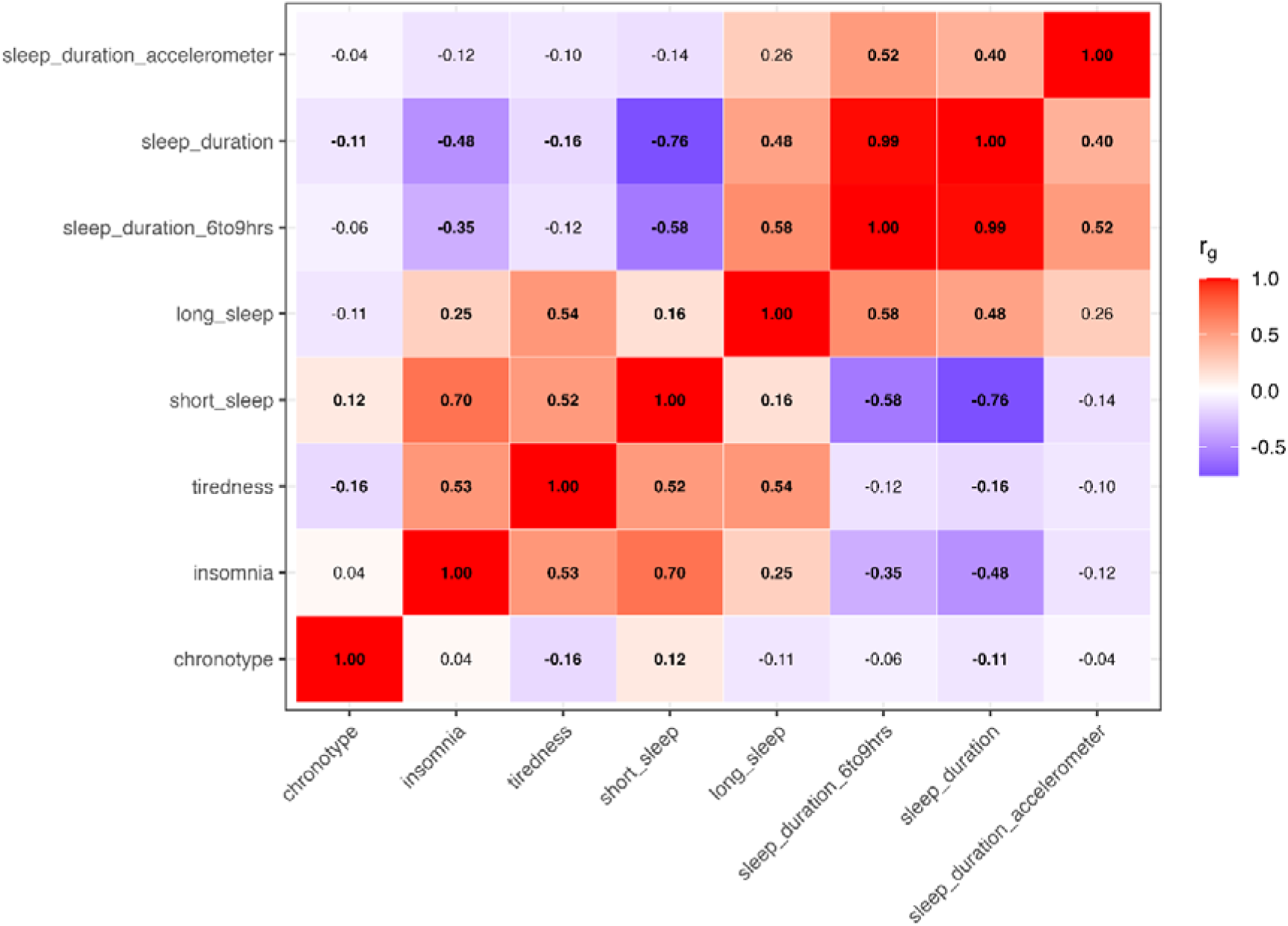
Genetic correlation between sleep traits

In addition, we conducted a sub-analysis of sleep duration within a ‘healthy’ range of six to nine hours. This analysis revealed a very similar trait to the overall sleep duration, with a genetic correlation of 0.99 (±0.007), p<1×10^-300^. We also considered the genetic correlation between self-reported sleep duration and accelerometer derived sleep duration (supplementary table 10) which revealed a correlation of rg=0.403 (±0.05), p=2.16 x10^-15^.

As observed when looking at sleep-related phenotypes, many traits relating to psychiatric conditions and cognitive abilities demonstrate a non-linear relationship, with a negative correlation to overall sleep duration and positive correlation with both short and long sleep (Figure 5).

**Figure 5.**
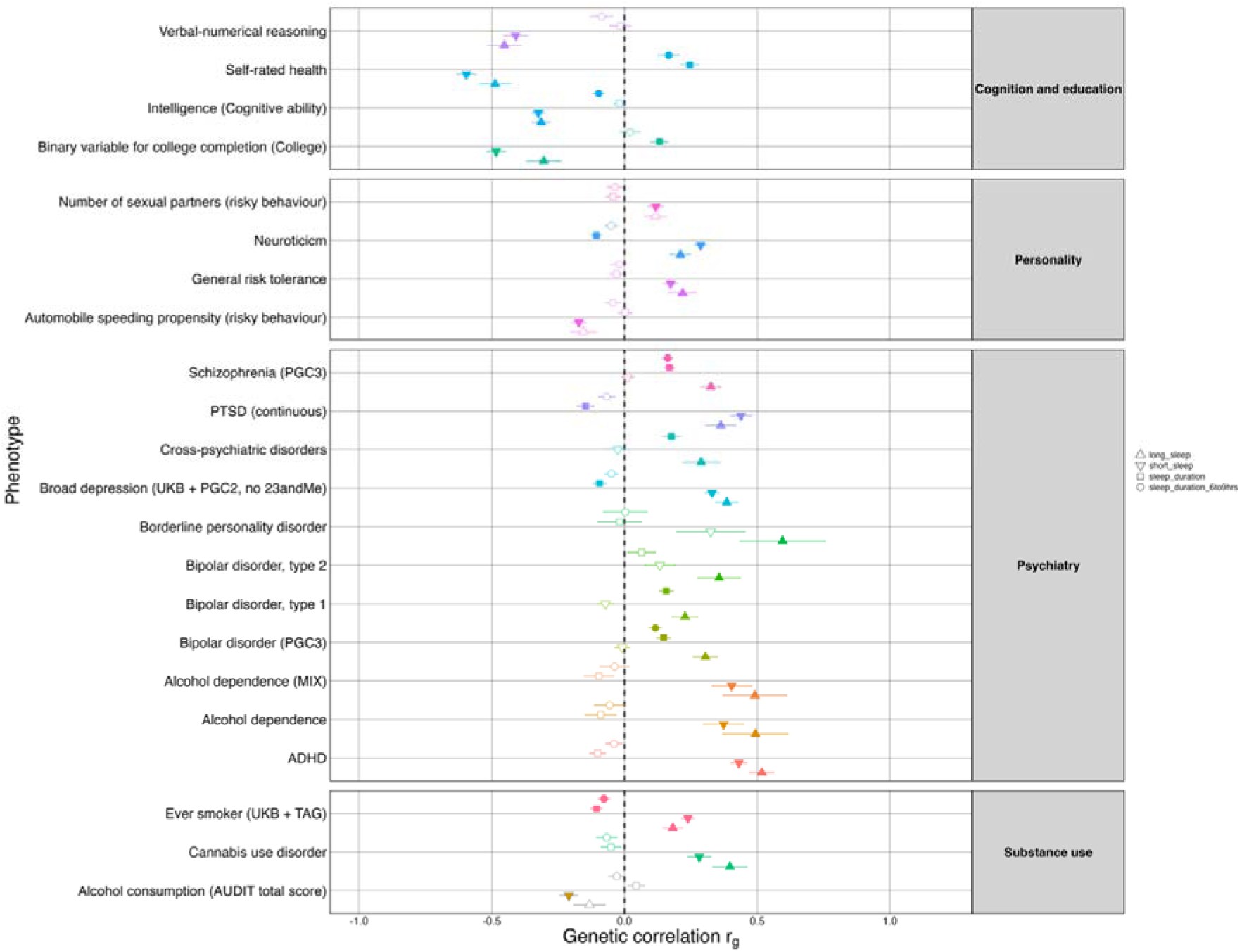
Genetic correlation between sleep duration and cognitive, psychiatric and personality-based phenotypes.

Local genetic correlation and cross-ancestry genetic correlation results are reported in the supplement.

### Genomic Structural Equation Modelling

We performed EFA and CFA of 15 genetically correlated traits. We considered multiple factor structures from one to six factors. We fitted models including either long and short sleep together, or quantitative sleep duration separately as models including all three related traits did not converge. We initially considered fifteen traits in total but could not fit a well-fitting l model inclusive of all these traits so the final models included either short/long sleep or continuous sleep duration, along with insomnia^8^, PTSD^41^, schizophrenia^42^, problematic alcohol use (PAU)^39^, alcohol consumption^35^, bipolar disorder^36^, cannabis use disorder (CUD)^37^, major depressive disorder (MDD)^38^, and Townsend deprivation index^44^.

The best fitting EFA model including short/long sleep (Figure 6, left panel) had five factors and the cumulative variance explained was 0.802, with SS loadings of 2.14, 2.11, 1.68, 1.61, 1.28 respectively. In this model, long sleep loaded with Townsend deprivation index, cannabis use disorder and problematic alcohol use, while short sleep loaded with insomnia only.

**Figure 6.**
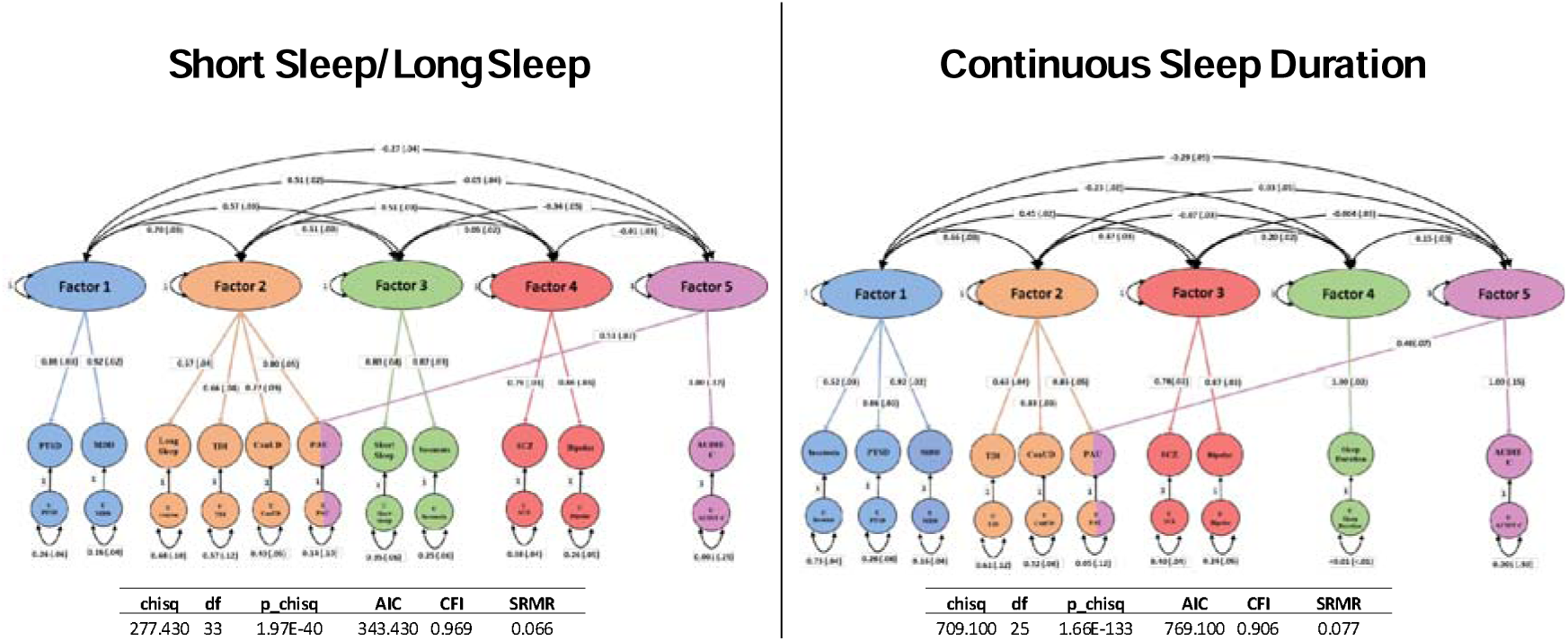
Genomic structural equation model

The best fitting EFA model including quantitative sleep (Figure 6, right panel) had five factors and the cumulative variance was 0.824, with SS loadings of 2.07, 2.05, 1.50, 1.33, 1.29 respectively. In this model, quantitative sleep duration was a separate factor with no co-loading traits (see supplementary table 11).

## Discussion

Sleep, and sleep duration are central to human health and illness. A better biological understanding of sleep, built on a knowledge of genetic factors that influence sleep duration, could create opportunities for improving sleep and therefore improving health and wellbeing. Prior work has focused primarily on sleep pathologies; here, we focus on how genetic variation relates to normal sleep duration. This study builds on a growing literature on the genetics of sleep duration, and identifies 234 associated loci, of which 129 have not been previously associated with any sleep trait, to our knowledge. The strongest association is rs2863957 on chromosome 2, within the *PAX8* gene, (±2.4 minutes, p=2.3×10^-66^). *PAX8* is now a well-replicated gene associated with overall sleep duration as well as short and long sleep^5,6,9,32^, so these findings support its role in modulating sleep duration. Of our novel findings, the variant with the largest sleep increasing association is rs144938821 on chromosome 2 (±2.6 minutes, p=8.9×10^-23^), which is an independent signal within *PAX8* and has previously been linked to insomnia. The locus with the greatest minute-wise effect on sleep is rs140736187 on chromosome 6 (±2.8 minutes, p=2.3×10^-8^), which maps to the *LINC01556*.

Though like most complex traits the average effect size was small (equating to less than one minute change in sleep duration), the cumulative effects of all risk loci equate to ±220 minutes. Most people will, of course, carry both decreasing and increasing sleep alleles, and future studies using these data to calculate polygenic risk scores in independent samples will give insight to how many sleep increasing and decreasing alleles people carry on average, and the extent to which this can predict differences in sleep patterns.

Our previous work on short and long sleep^5^ demonstrated a modest, positive genetic correlation between genetic liability for these two extremes of sleep duration. Here, we show that overall sleep duration is positively correlated to long sleep (rg=0.48, sd=0.03, p=1.7 x10^-47^) and negatively correlated to short sleep (rg=-0.76, sd=0.01, p<1×10^-300^). We conducted an additional GWAS of sleep duration within the normal or healthy range of six to nine hours. This trait, which excludes a total of 63,016 extreme short/long sleepers, has a genetic correlation of >0.99 with overall sleep. When comparing this truncated healthy range sleep GWAS to long sleep we see a correlation of rg=0.58 (sd=0.05, p=3.42e-27), and to short sleep rg=-0.58 (sd=0.03, p=1.48 x10^-91^). Taken together, this suggests that a GWAS of overall sleep duration (1-23 hours) largely captures the genetic basis of healthy sleep. We additionally show several examples of phenotypes with a negative genetic correlation to overall sleep duration, but a positive genetic correlation to both short and long sleep (figure 5). This supports our previous assertion^5^ that short and long sleep should be considered as separate traits and not extremes of the same continuum.

We also compared our findings to other measures of sleep. Outside of sleep duration, the most widely studied sleep traits are insomnia and chronotype (measured as morningness or morning chronotype). Insomnia and chronotype are themselves not genetically correlated (rg=0.04, sd=0.02, p=0.09). As figure 4 shows, we find a negative correlation between sleep duration and insomnia, and a more modest negative corelation between sleep duration and chronotype (insomnia: rg=-0.48, sd=0.02, p=9.9e-86; chronotype: rg=-0.11, sd=0.02, p=2.13 x10^-6^). The genetics of chronotype have been described in a large GWAS of almost 700,000 subjects which identified several loci in genes involved in circadian regulation^46^. These genes, including *CRY1* and *PER2/3*, are not observed in GWAS of sleep duration or insomnia. This indicates that the biological pathways influencing sleep timing are largely distinct from those that regulate sleep duration and sleep quality.

We further interrogated the shared and independent genetic architecture between sleep and various other traits using gSEM^34^, focusing on traits positively correlated with both long and short sleep duration. We generated two five-factor models, one including short and long sleep duration and another including overall sleep duration. When we included short and long sleep duration, we observed factor loading that reflected clinical expectations, with short sleep loading with insomnia and long sleep loading with problematic substance use outcomes known to impact sleep quality. When we included overall sleep duration, it loaded on its own factor with relatively low correlations to other factors demonstrating that this trait reflects healthier range sleep duration and is largely independent of the psychiatric traits included in this model.

Self-reported sleep duration is a simple and replicable measure, making it a widely used metric of sleep quality. Previous work has compared self-reported sleep duration with an objective measure of sleep patterns, using accelerometer-derived data from the UKB^6^. They found that the lead SNPs for sleep duration associated with the accelerometer derived sleep duration estimates, as well as with short and long sleep, sleep efficiency and number of sleep bouts, but not with measures of sleep timing. Here, we demonstrate a positive correlation of 0.4 between self-reported sleep duration measure and accelerometer derived sleep duration^6^. This is much lower than the correlation estimates between self-reported sleep duration and being a self-described ‘undersleeper’ or ‘oversleeper’, and lower even than the correlation between self-reported sleep duration and insomnia. This suggests that our data might be better understood as describing the genetic basis of perceived rest, not necessarily an accurate representation of actual hours slept, incorporating the functional impact of the individual’s sleep. This bears consideration particularly when looking at the correlation estimates seen between self-reported sleep duration and various psychiatric traits. Future studies using wearable technology will be valuable in investigating this further.

In the second model, short sleep loads with insomnia while long sleep loads with Townsend deprivation index, cannabis use disorder, alcohol consumption and problematic alcohol use (which co-loads with AUDIT-C, a measure of alcohol consumption). The correlation between these two factors is 0.51. Epidemiological studies demonstrate that both cannabis and alcohol are frequently used by people to help sleep onset, but also that prolonged use of both substances is associated with sleep disturbance^47,48^.

We investigated how the genome wide results relate to specific cell types using polygenic regression analysis with reference to a scRNA-seq dataset that included 10 biological systems and found significant associations in each of these systems, reflecting the biologically pervasive importance of sleep. The lead findings implicated lung, liver, immune, brain, and eye cell types, in that order. The importance of the brain and eye (e.g. as an input for circadian regulation) to sleep regulation are well known, as is the impact of sleep on immune function. Less well recognized are an association of sleep with lung and liver function, although both long and short sleep duration are associated with lower lung function and higher risk of asthma, and long sleep duration with computed tomographic lung abnormalities^49,50^, while an association of sleep duration with metabolic dysfunction–associated steatotic liver disease (previously called nonalcoholic fatty liver disease) is variously reported but inconclusive^51^

The findings reported here should be considered in the context of several limitations. Firstly, as described previously we rely on self-reported data to define ‘sleep duration’ meaning this may, to some extent, be capturing levels of perceived rest rather than exact hours slept. Additionally, the question in both UKB and MVP that was used to derive this data asked for typical hours slept in a 24-hour period. This could mean we are combining people who get a single block of sleep during the night with those who have interrupted low-quality sleep and then compensate with naps during the day. This should be considered when interpreting the relationships between sleep duration and other phenotypes. Secondly, sleep is a highly complex trait impacted by multiple environmental factors. We conducted sensitivity analyses in our previous work to show that the genetic architecture of sleep appears to be robust to some of these factors (e.g. shift work, sex)^5^, but we have not tested the impact of other factors known to impact sleep such as caffeine consumption, daylight exposure, or substance use. Finally, although we included the available non-EUR data in both UKB and MVP, the majority of participants in both cohorts are of European genetic ancestry. As such, these findings cannot be considered generalisable to the global population and should be repeated in large, diverse samples as these become available.

Overall, we highlight several novel SNPs and genes associated with quantitative sleep duration which give new avenues for future research on the biology underlying sleep. This work adds to a growing literature on the genetic basis of sleep which, given the wealth of evidence supporting the role of healthy sleep patterns in supporting overall wellbeing, has significant public health importance. However, we argue that sleep duration as a continuous trait is less relevant to the relationship between poor sleep and negative health consequences than are measures of extreme long and short sleep duration; and more relevant to the understanding of health.

## Data Availability

All data produced in the present study will be available upon reasonable request to the authors following publication in a peer reviewed journal.

## Acknowledgements

This research has been conducted using the UK Biobank (www.ukbiobank.ac.uk) under application numbers 82087 (PI: Dr Jonathan Coleman). This research is also based on data from the Million Veteran Programme, Office of Research and Development, Veterans Health Administration. This publication does not represent the views of the Department of Veteran Affairs or the United States Government. This work is supported by funding from the UK Medical Research Council and the US Department of Veteran Affairs (CSP575b (NCT02256644) and MERIT (I01CX001849) grants) (both JG and MBS). JG and MBS report support from NIMH R01MH133728-01. D.F.L. was supported by a NARSAD Young Investigator Award from the Brain & Behavior Research Foundation and a Career Development Award from the Veterans Health Administration Office of Research and Development (Grant IK2BX005058) and is Aimee Mann Fellow of Psychiatric Genetics at Yale. J.D.D. was supported by the National Institute on Alcohol Abuse and Alcoholism (NIAAA) T32 AA028259.

The authors thank all the volunteers who participated in the UK Biobank and the Million Veteran Programme. We gratefully acknowledge all the studies and databases that made GWAS summary data available.

