## supplementary material for "Multi-ancestry genetic architecture of sleep duration and its relationship to other sleep and psychiatric phenotypes"

S-Figure 1. Reported sleep duration in hours in the UK Biobank (UKB) and Million Veteran Program (MVP)

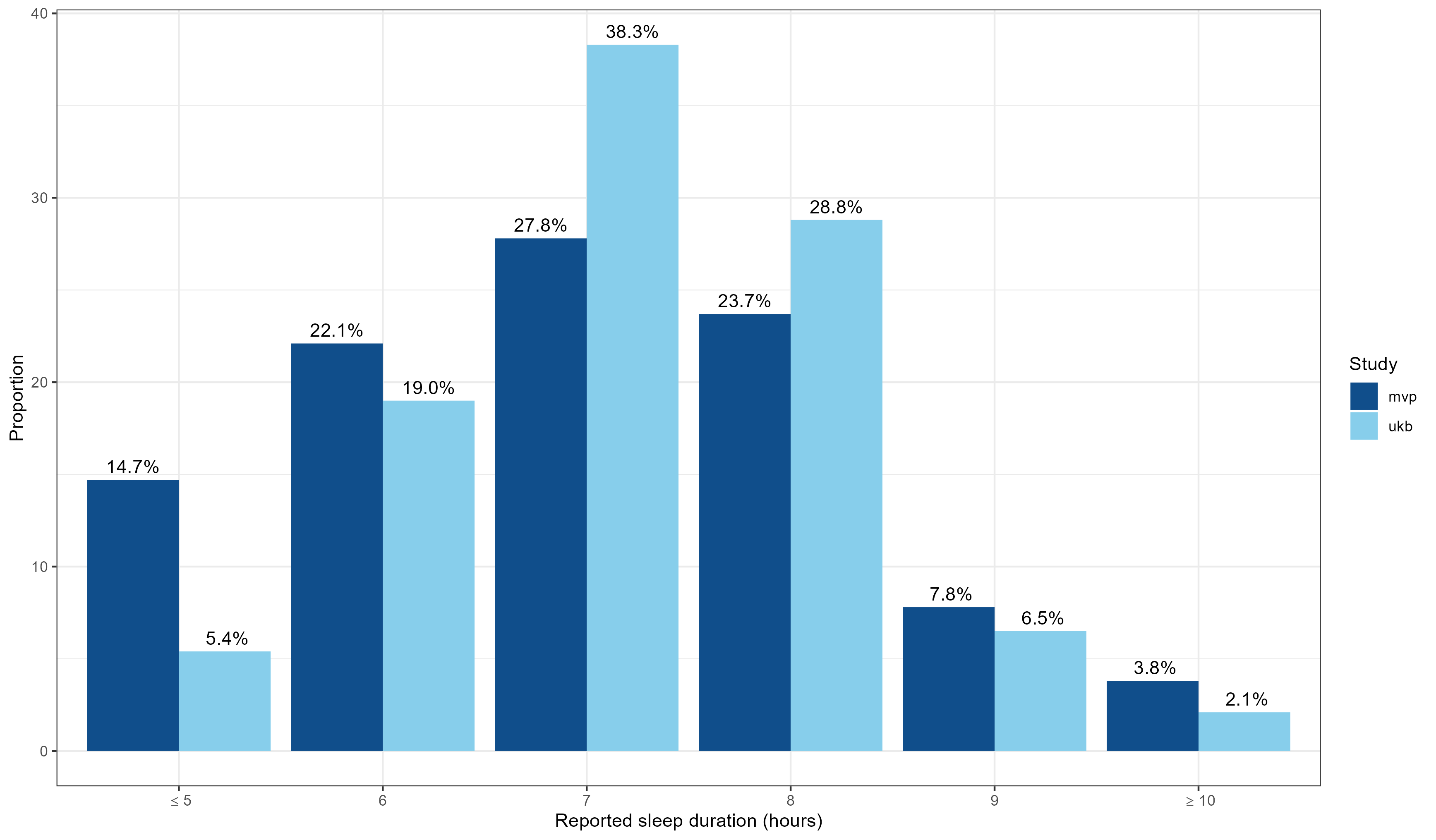

S-Figure 2. QQ-plot for cross-population meta-analysis of quantitative sleep duration in the UKB and MVP samples.

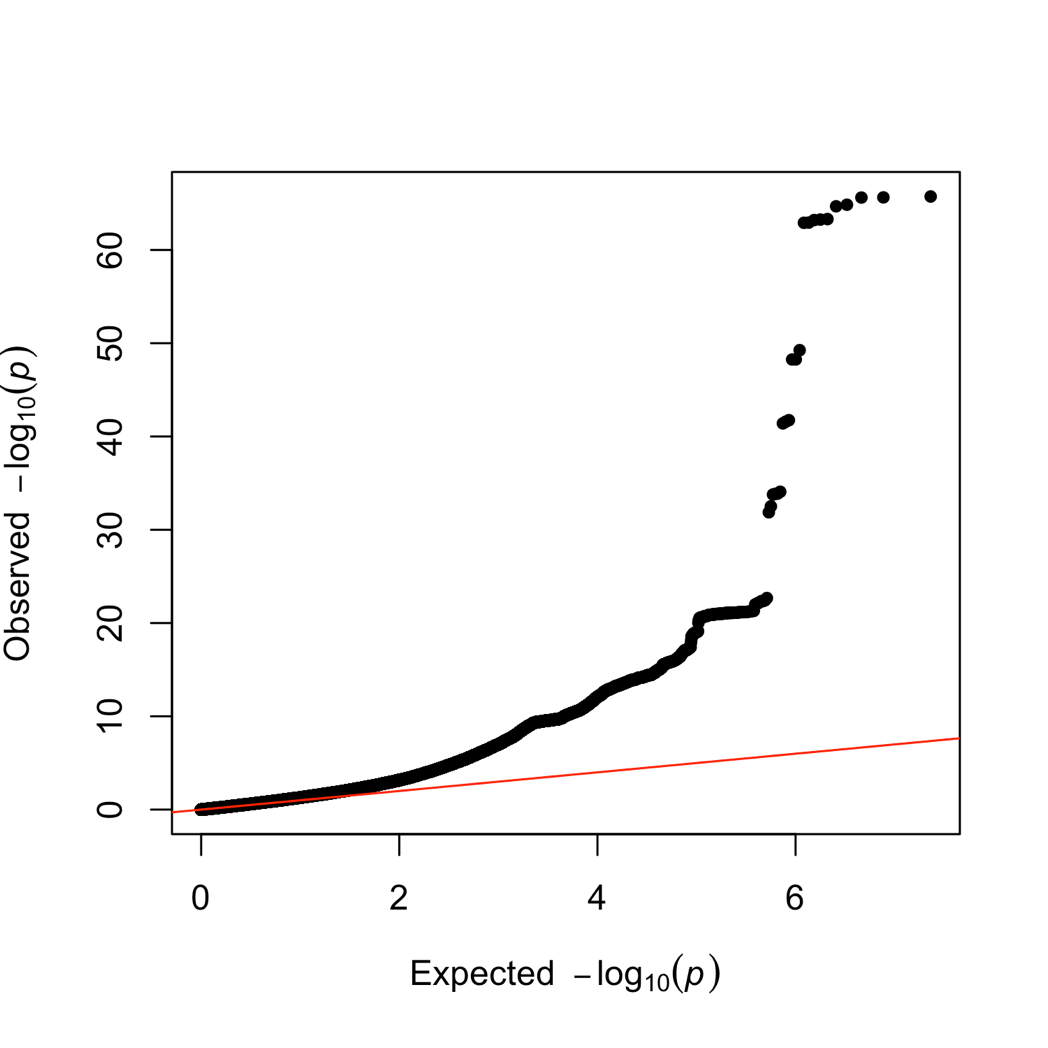

S-Figure 3. Manhattan plot showing the results for the EUR meta-analysis of quantitative sleep duration in the UKB and MVP samples.

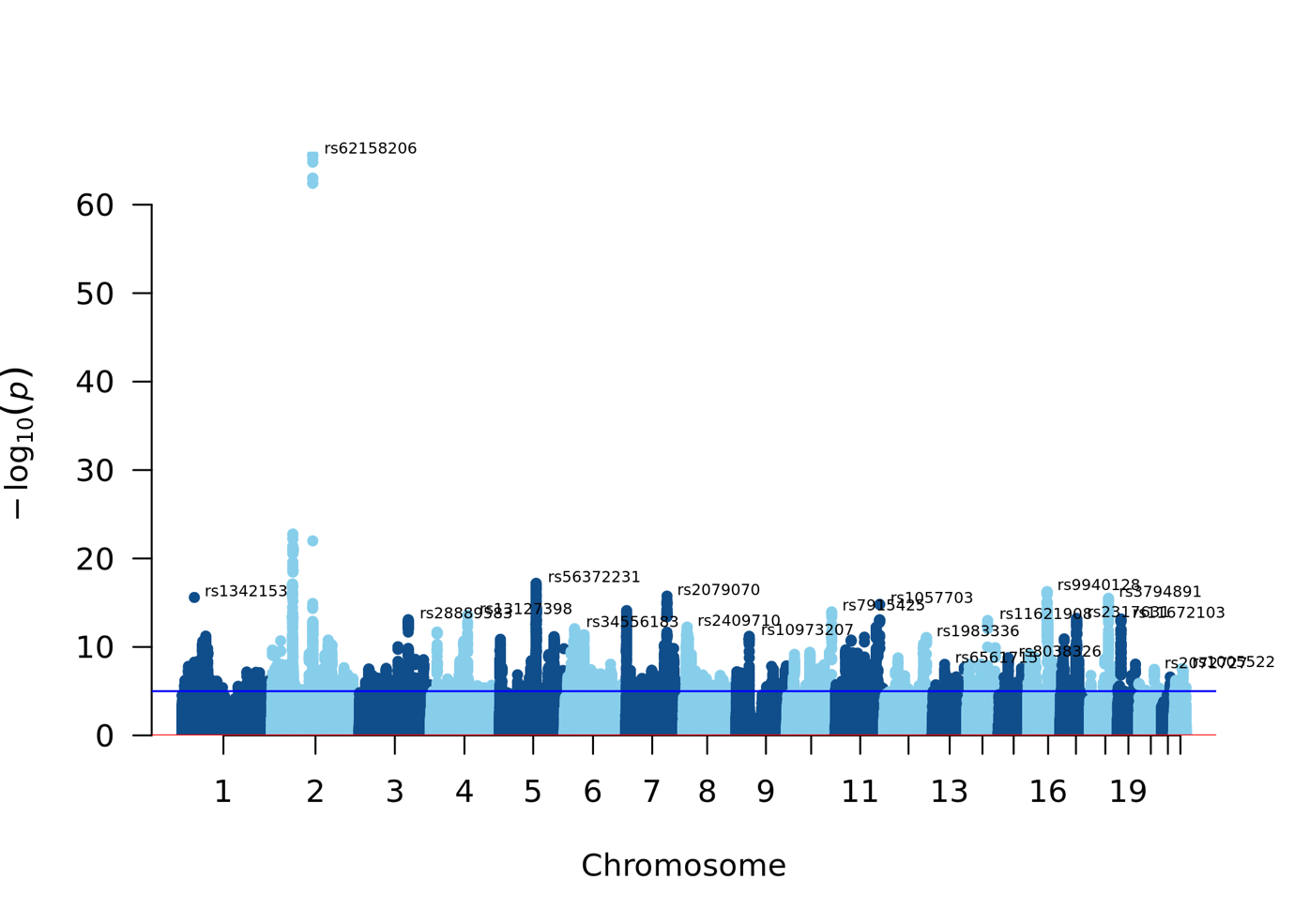

S-Figure 4. QQ-plot for EUR meta-analysis of quantitative sleep duration in the UKB and MVP samples.

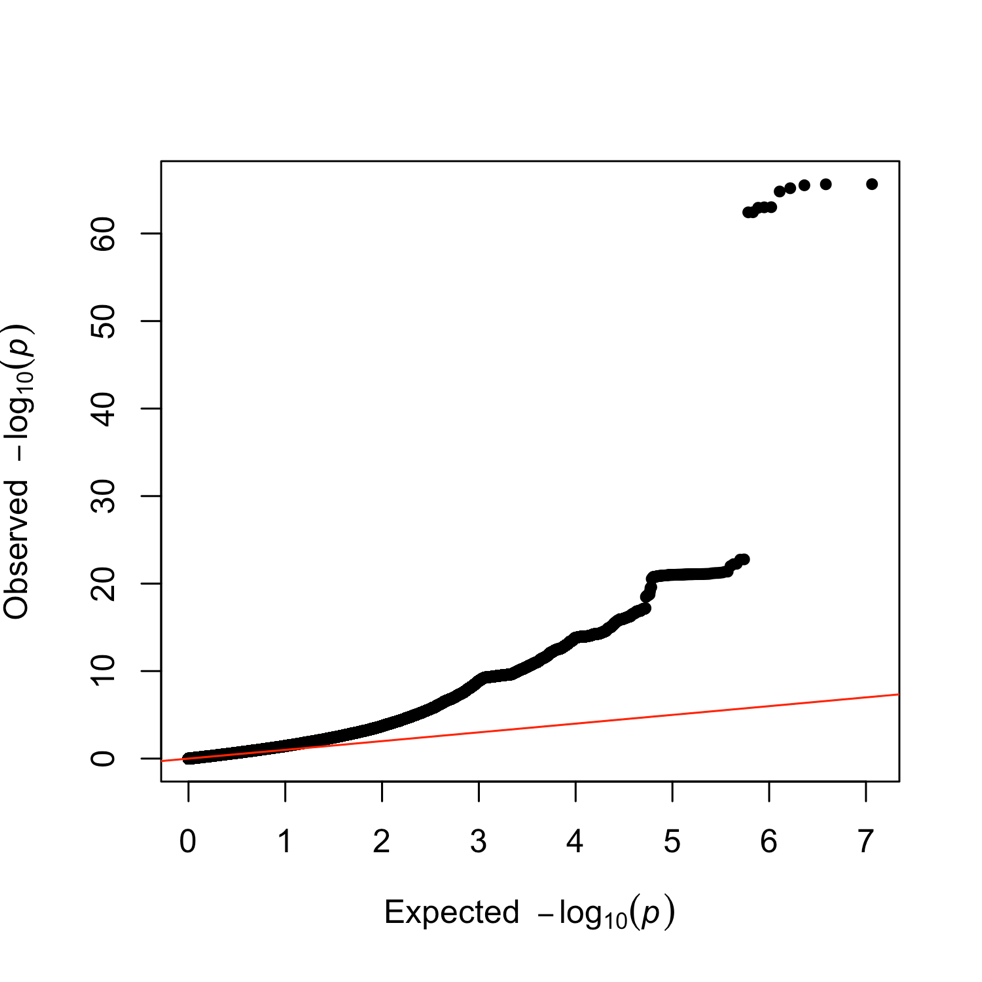

S-Figure 5. Gene-based Manhattan plot showing the results for the EUR meta-analysis of quantitative sleep duration in the UKB and MVP samples.
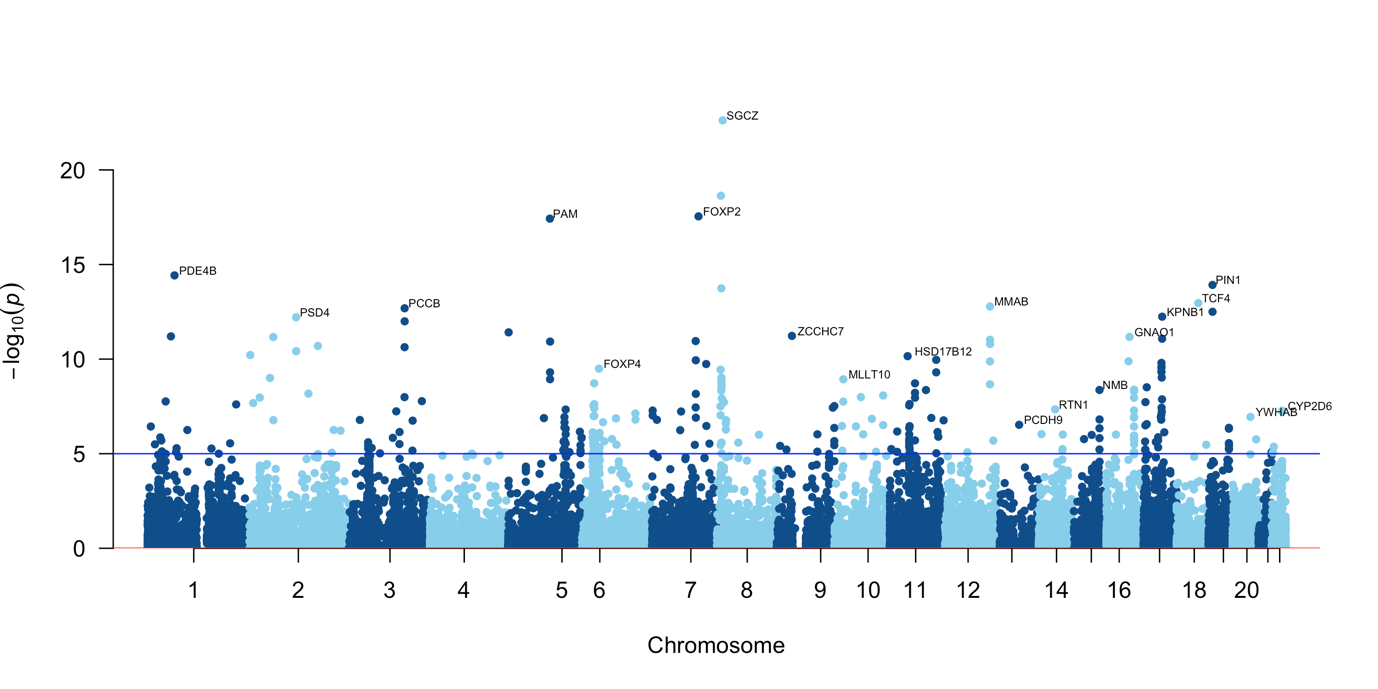

S-Figure 6. Manhattan plot showing the results for the AFR meta-analysis of quantitative sleep duration in the UKB and MVP samples.

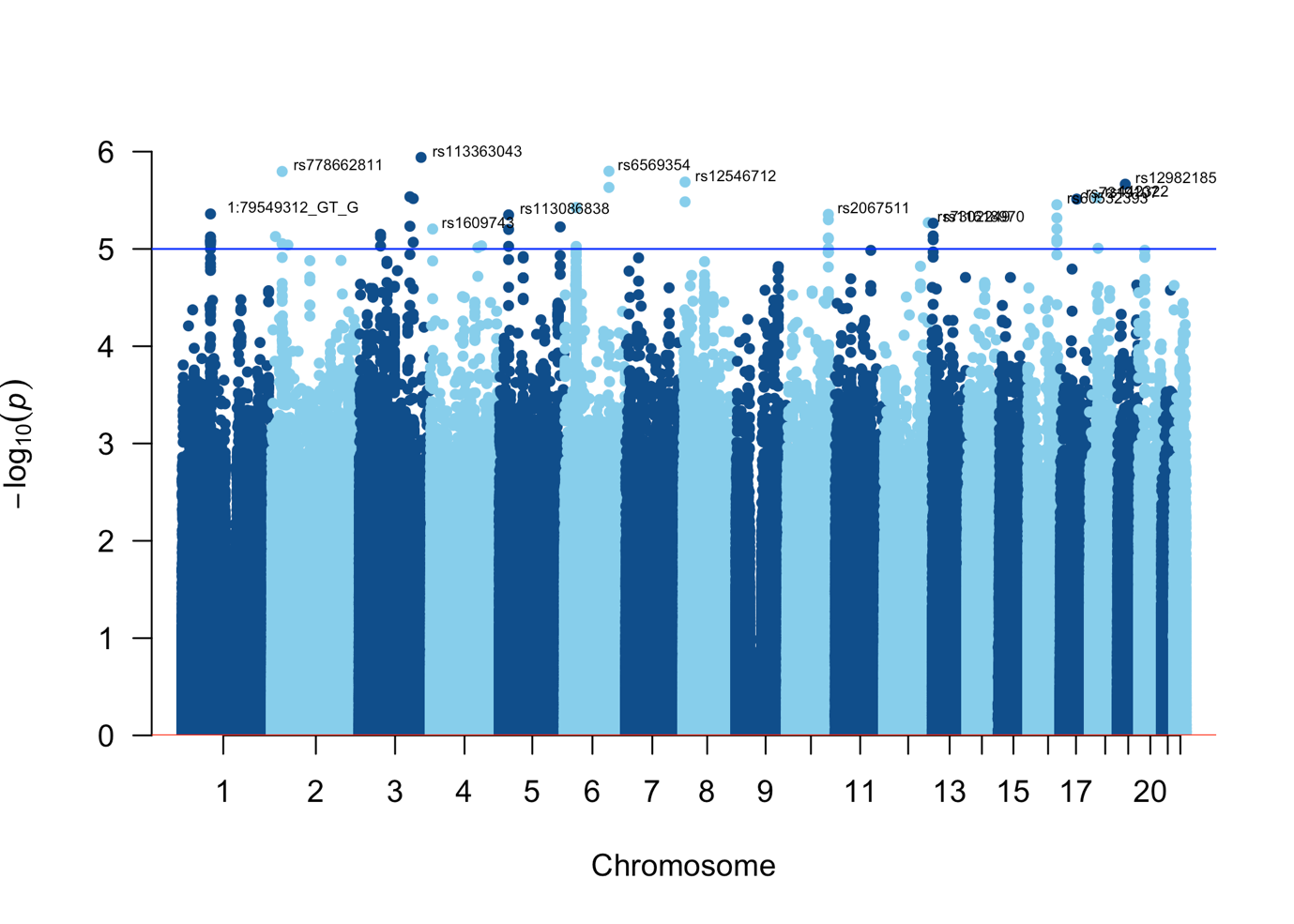

S-Figure 7. QQ-plot for AFR meta-analysis of quantitative sleep duration in the UKB and MVP samples.

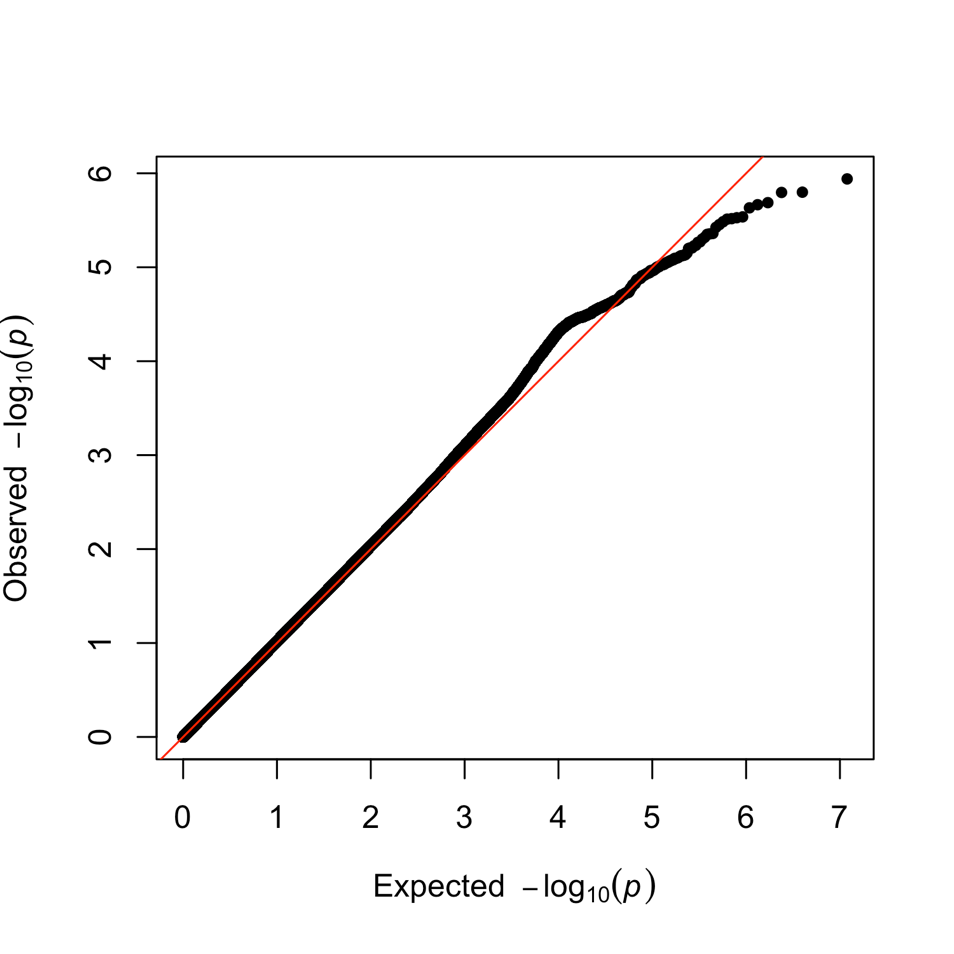

S-Figure 8. Gene-based Manhattan plot showing the results for the AFR meta-analysis of quantitative sleep duration in the UKB and MVP samples.

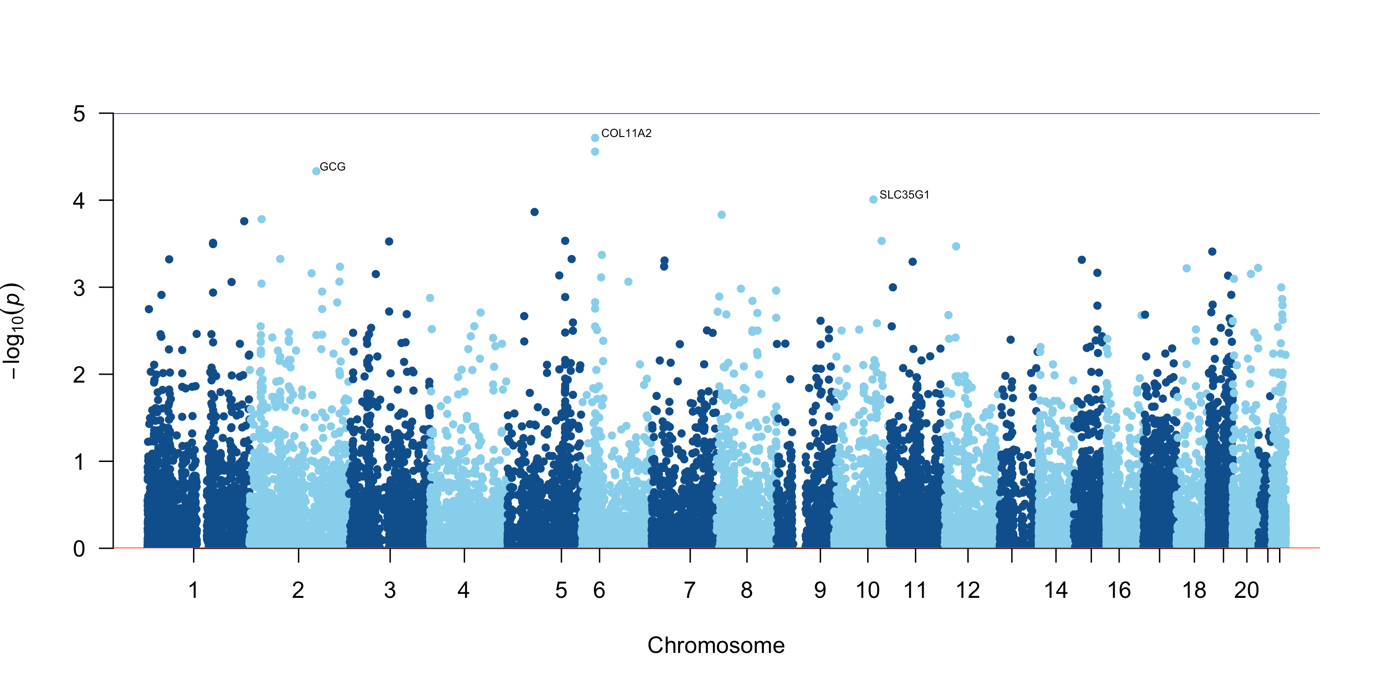

S-Figure 9. Pathway analysis results from PASCAL analysis reveals three pathways reaching a Bonferroni-corrected significance threshold of p<4.6e-5.

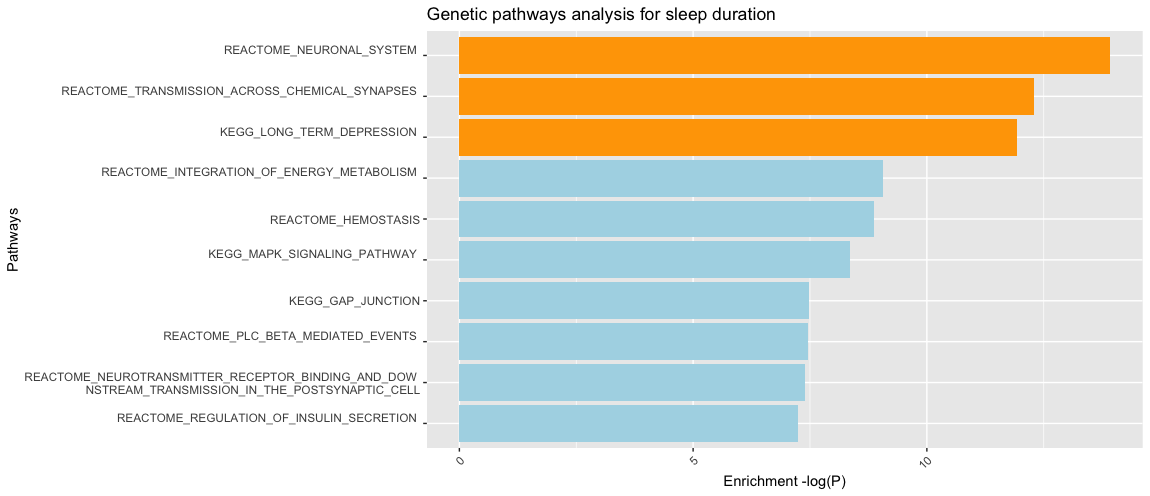

S-Figure 10. TWAS Z-score associations with brain tissues

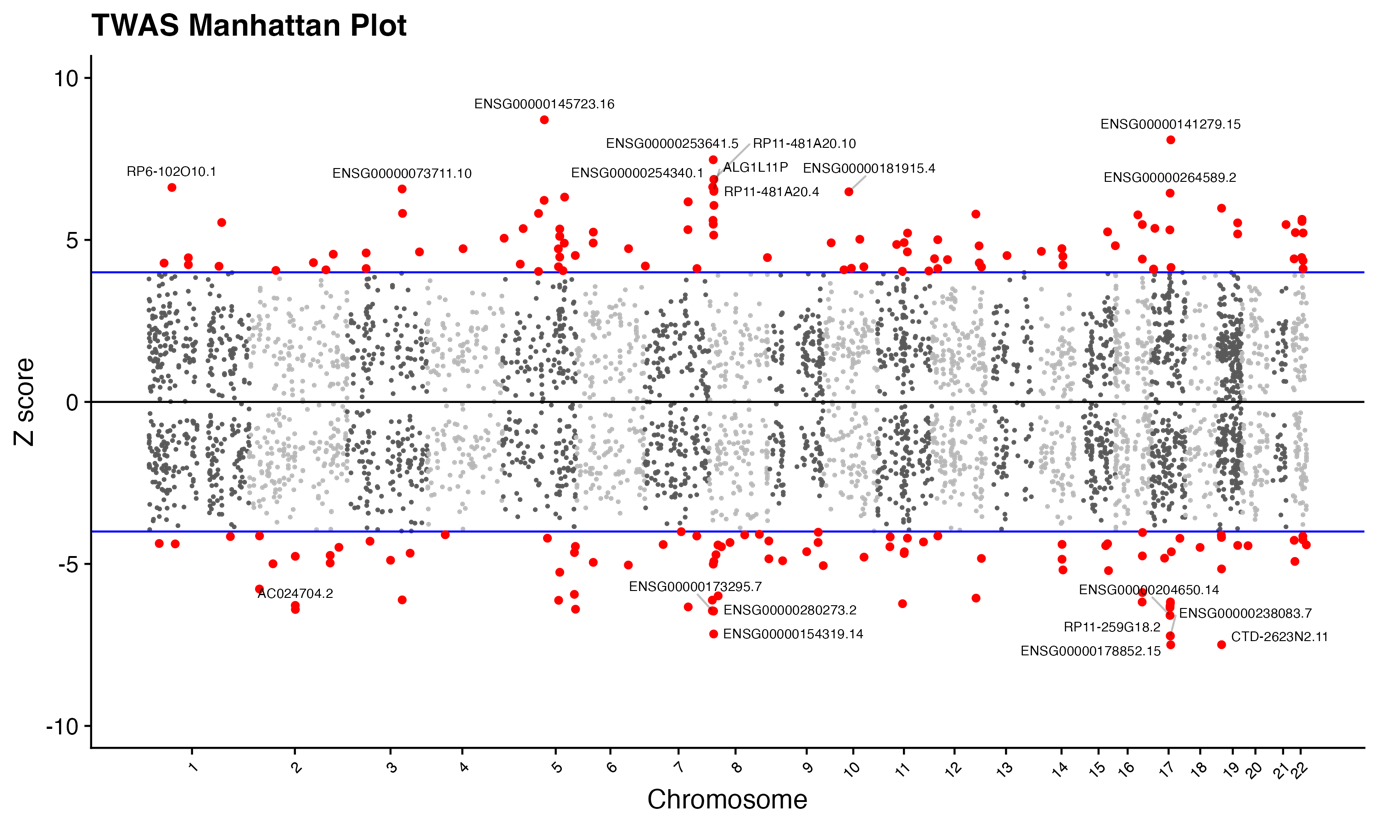

S-Figure 11. TWAS Z-score associations with non-brain tissues

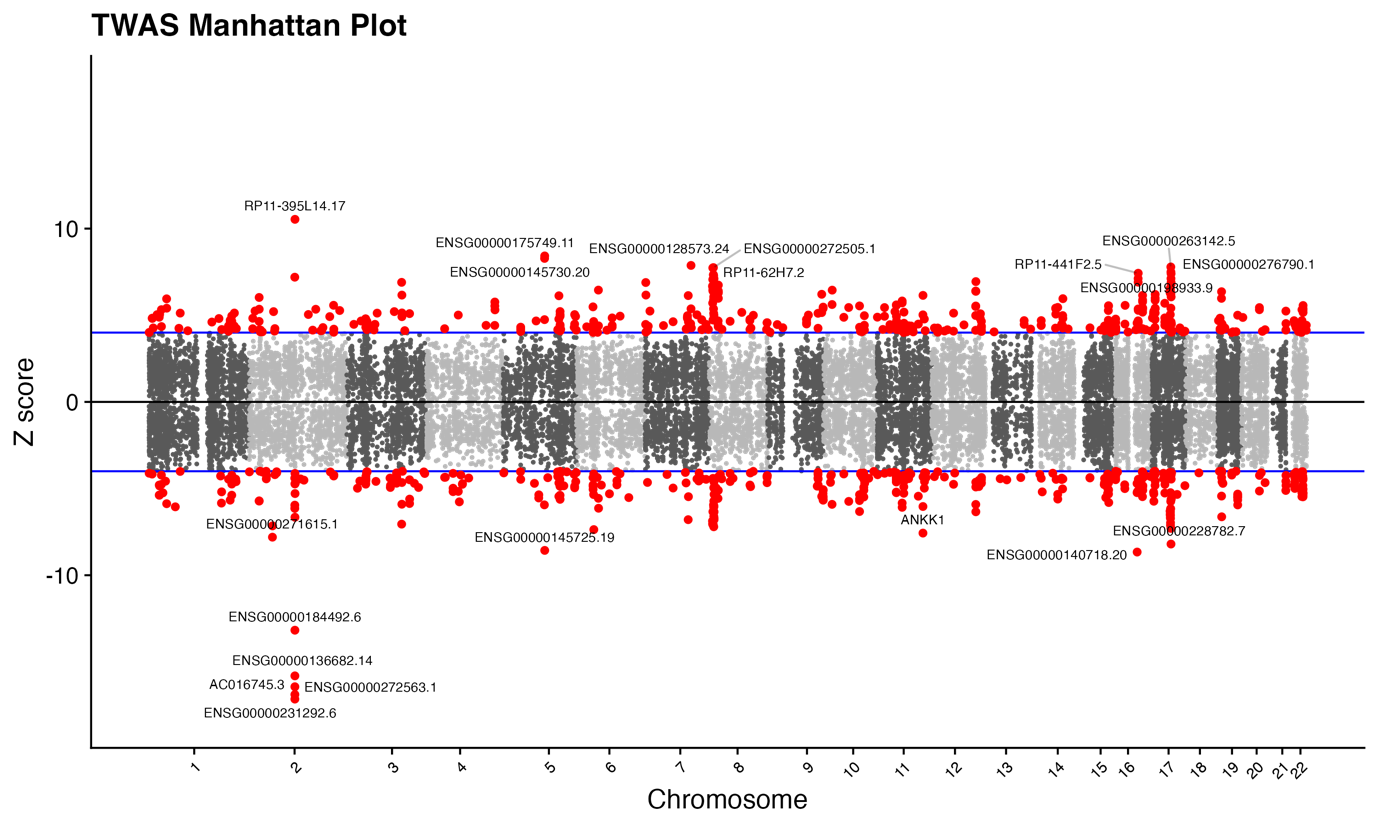

S-Figure 12. Single-cell analysis – eye

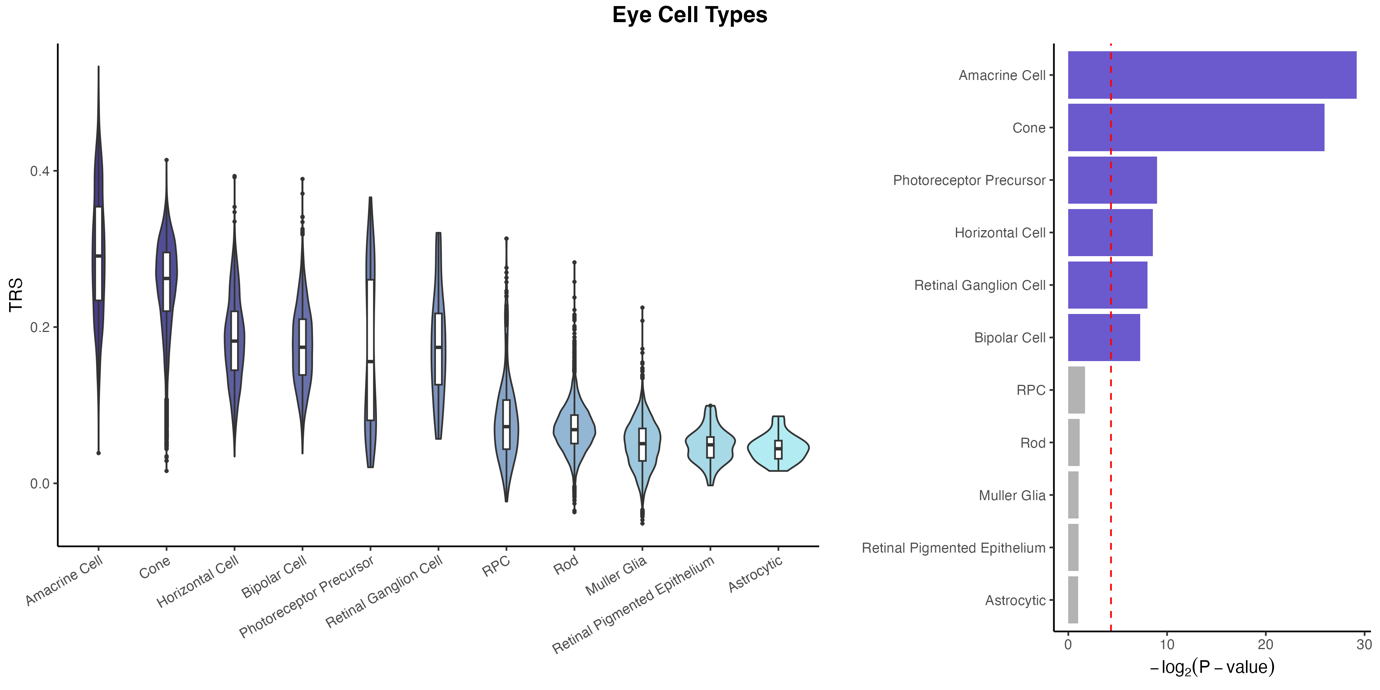

S-Figure 13. Single-cell analysis – heart

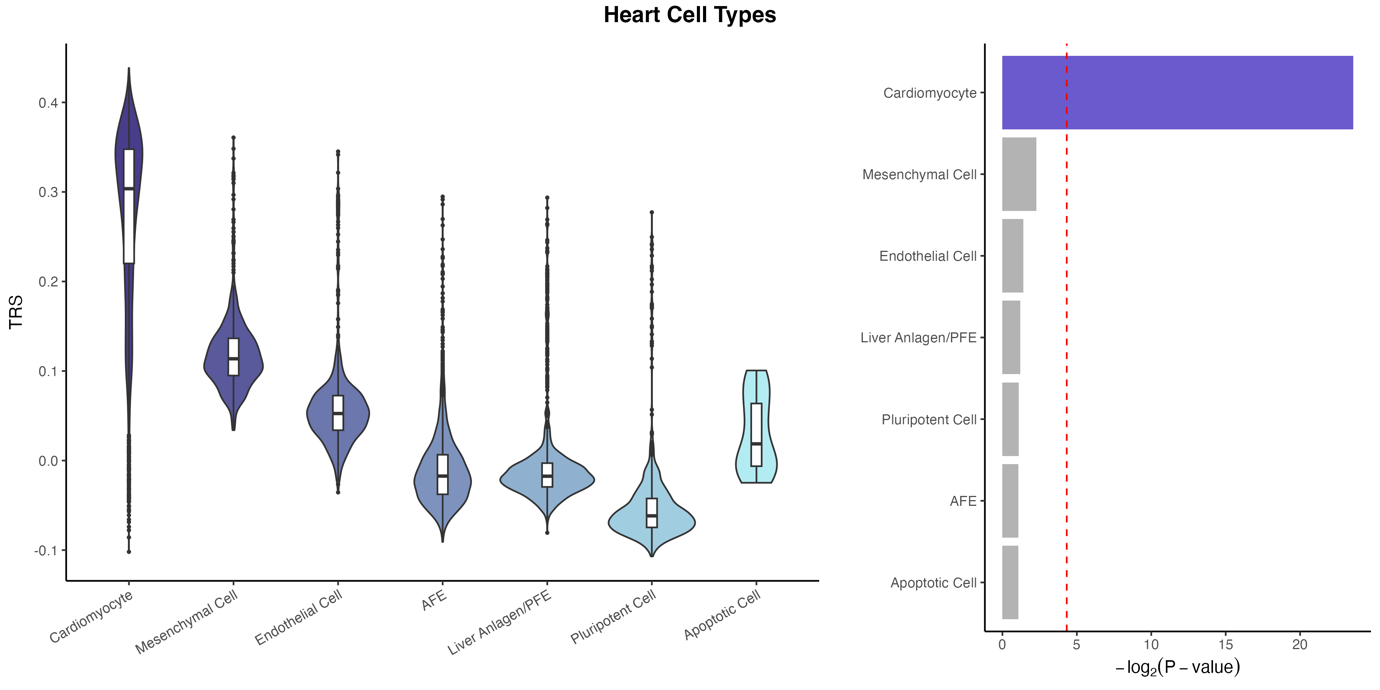

S-Figure 14. Single-cell analysis – immune system

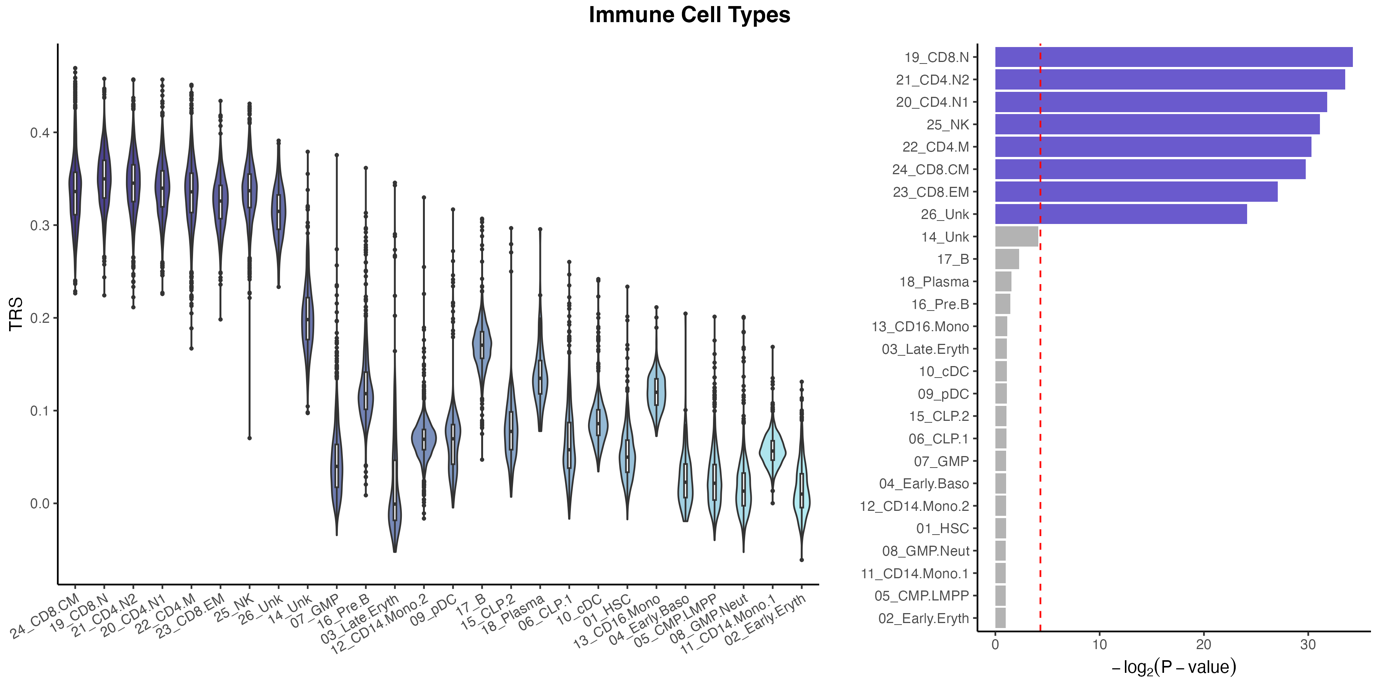

S-Figure 15. Single-cell analysis – intestine

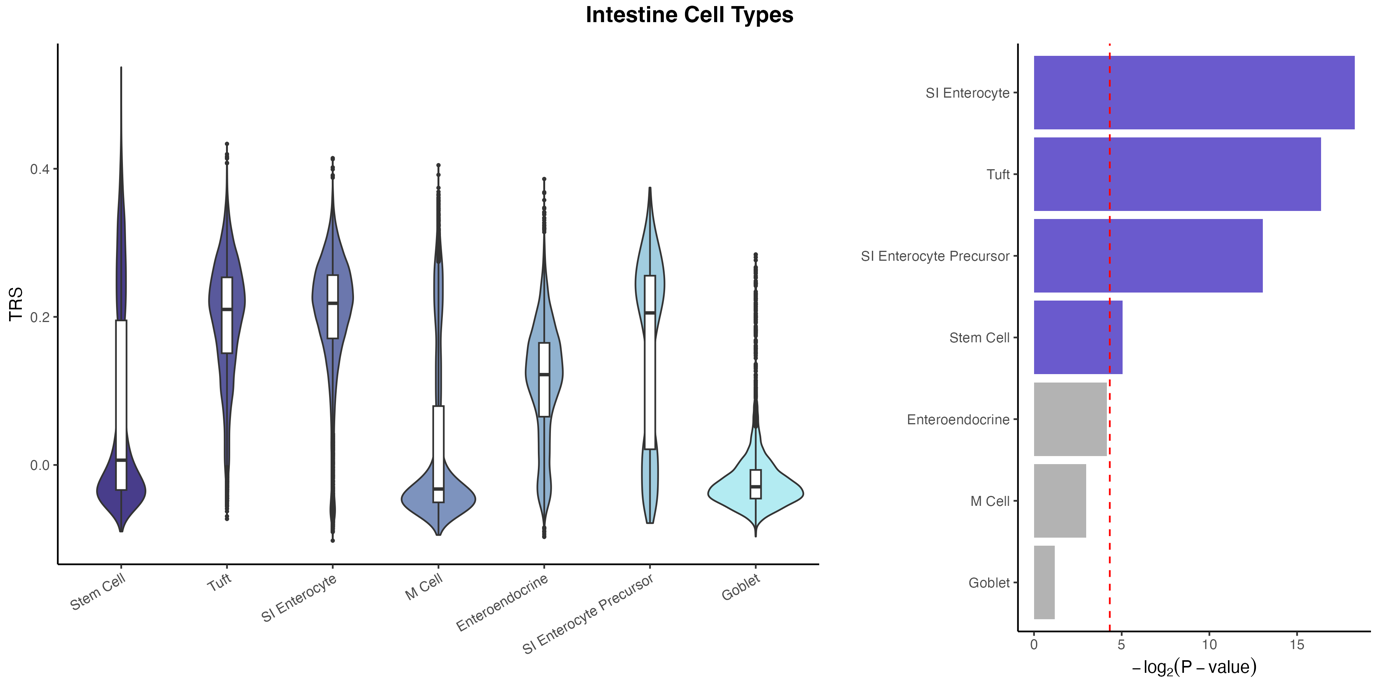

S-Figure 16. Single-cell analysis – kidney

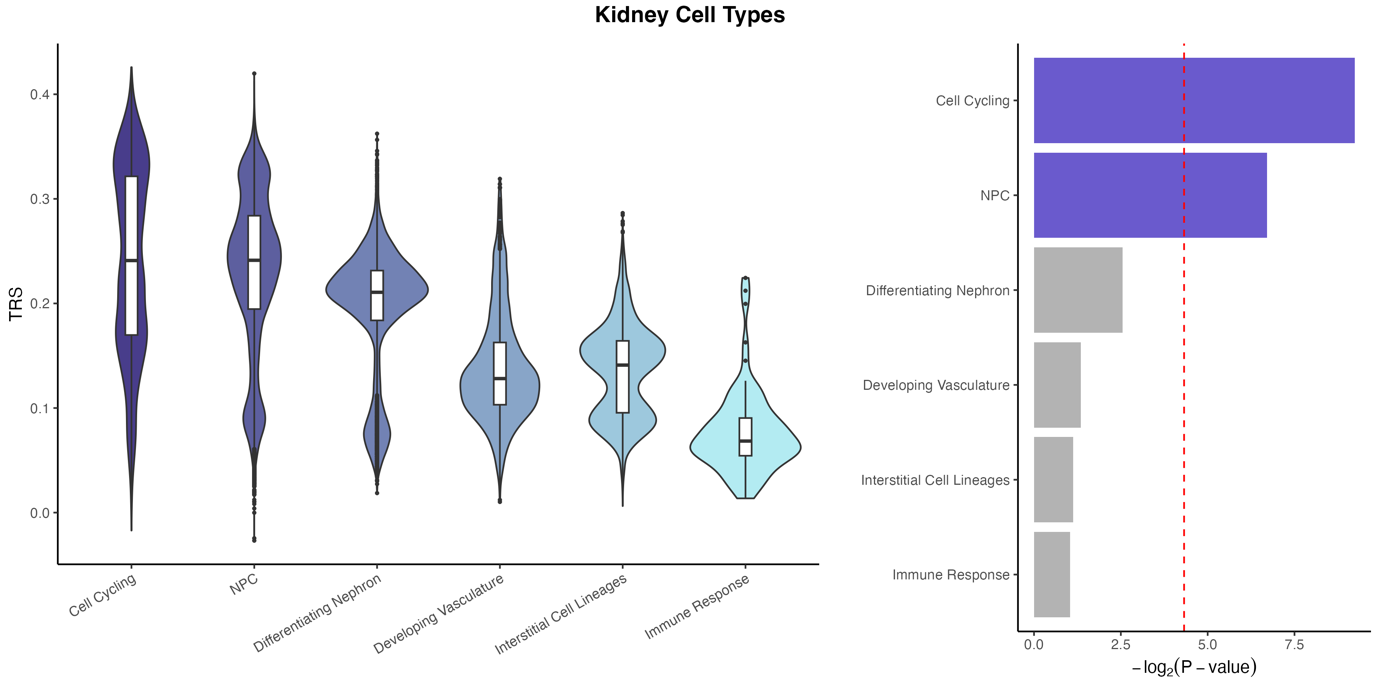

S-Figure 17. Single-cell analysis – liver

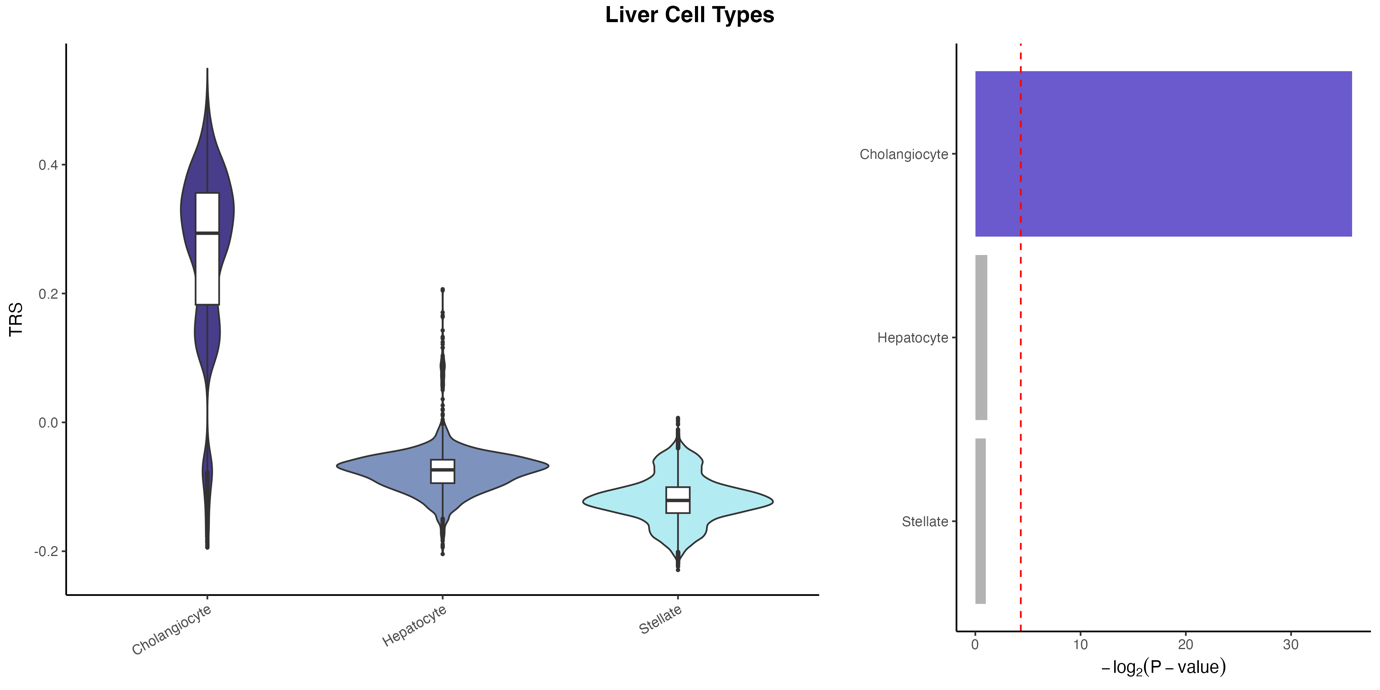

S-Figure 18. Single-cell analysis – pancreas

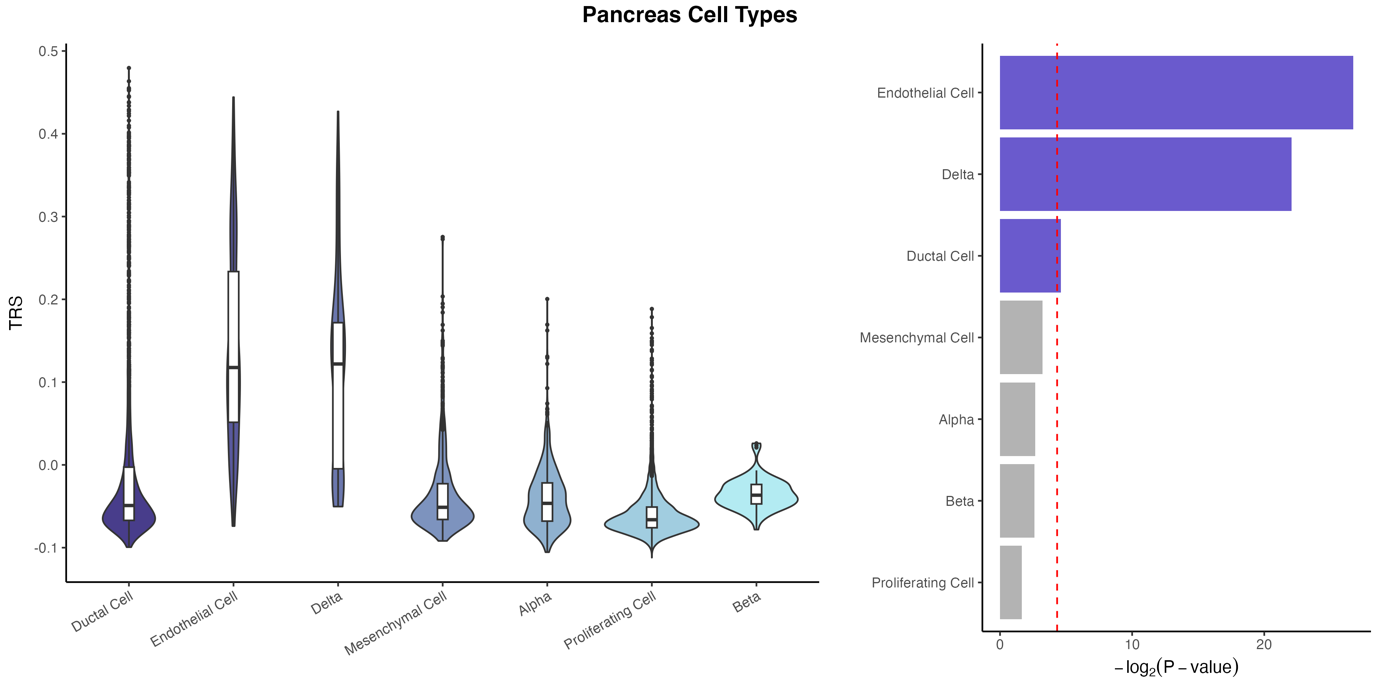

S-Figure 19. Single-cell analysis – skin

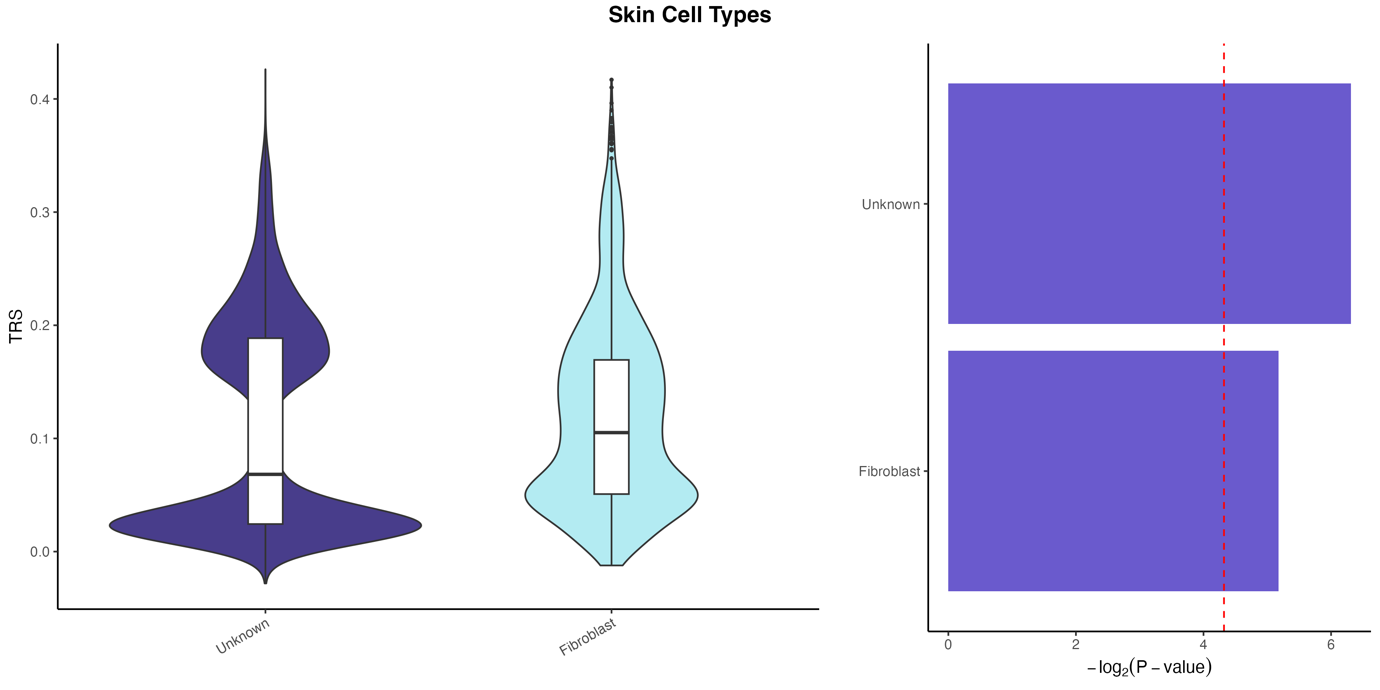

S-Figure 20. Single-cell analysis – brain

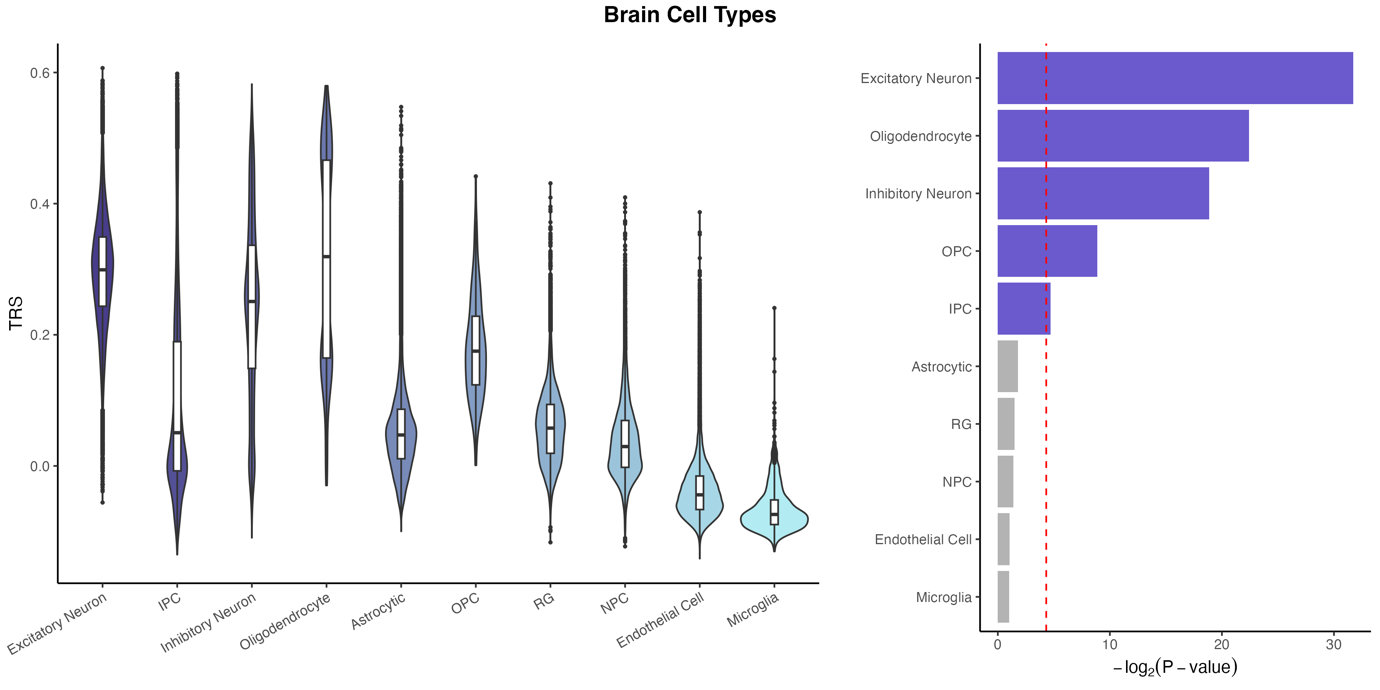

S-Figure 21. Single-cell analysis – lung

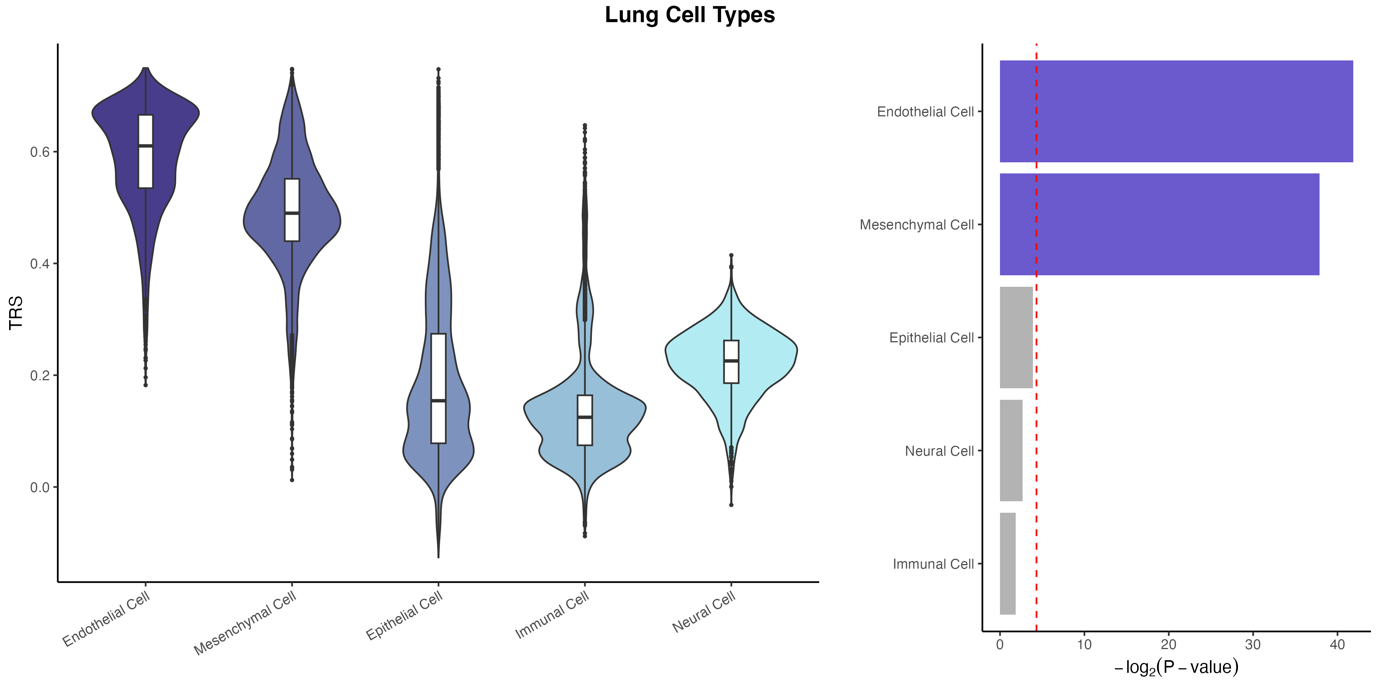
